# Medical students’ perceptions towards artificial intelligence in education and practice: A multinational, multicenter cross-sectional study

**DOI:** 10.1101/2023.12.09.23299744

**Authors:** Felix Busch, Lena Hoffmann, Daniel Truhn, Esteban Ortiz-Prado, Marcus R. Makowski, Keno K. Bressem, Lisa C. Adams, COMFORT Consortium

## Abstract

**Background:** Artificial intelligence (AI) is anticipated to fundamentally change the educational and professional landscape for the next generation of physicians, but its successful integration depends on the global perspectives of all stakeholders. Previous medical student surveys were limited by small sample sizes or geographic constraints, hindering a global comparison of perceptions. This study aims to explore current medical students’ attitudes towards AI in medical education and the profession on a broad, international scale and to examine regional differences in perspectives.

**Methods and Findings:** This international multicenter cross-sectional study developed and validated an anonymous online survey of 15 multiple-choice items to assess medical, dentistry, and veterinary students’ AI knowledge and attitudes toward the utilization of AI in healthcare, the current state of AI education, and regional differences in perspectives. Between April and October 2023, 4,313 medical, 205 dentistry, and 78 veterinary students from 192 faculties in 48 countries responded to the survey (average response rate: 0.2%, standard deviation: 0.4%). Most participants studied in European countries (N=2,350), followed by North/South America (N=1,070) and Asia (N=944). Students expressed predominantly positive attitudes towards the use of AI in healthcare (67.6%, N=3,091) and the desire for more AI teaching in their curricula (76.1%, N=3,474). However, they reported limited general knowledge of AI (75.3%, N=3,451), the absence of AI-related courses (76.3%, N=3,497), and felt inadequately prepared to use AI in their future careers (57.9%, N=2,652). The subgroup analyses revealed regional differences in perceptions, although predominantly with small effect sizes. The main limitations include the low response rate per institution, which was calculated on total enrollment across all degree programs, and the risk of selection bias.

**Conclusions:** This study highlights the favorable perceptions of international medical students towards incorporating AI in healthcare practice while emphasizing the importance of integrating AI teaching into medical education.

**Graphical abstract:** 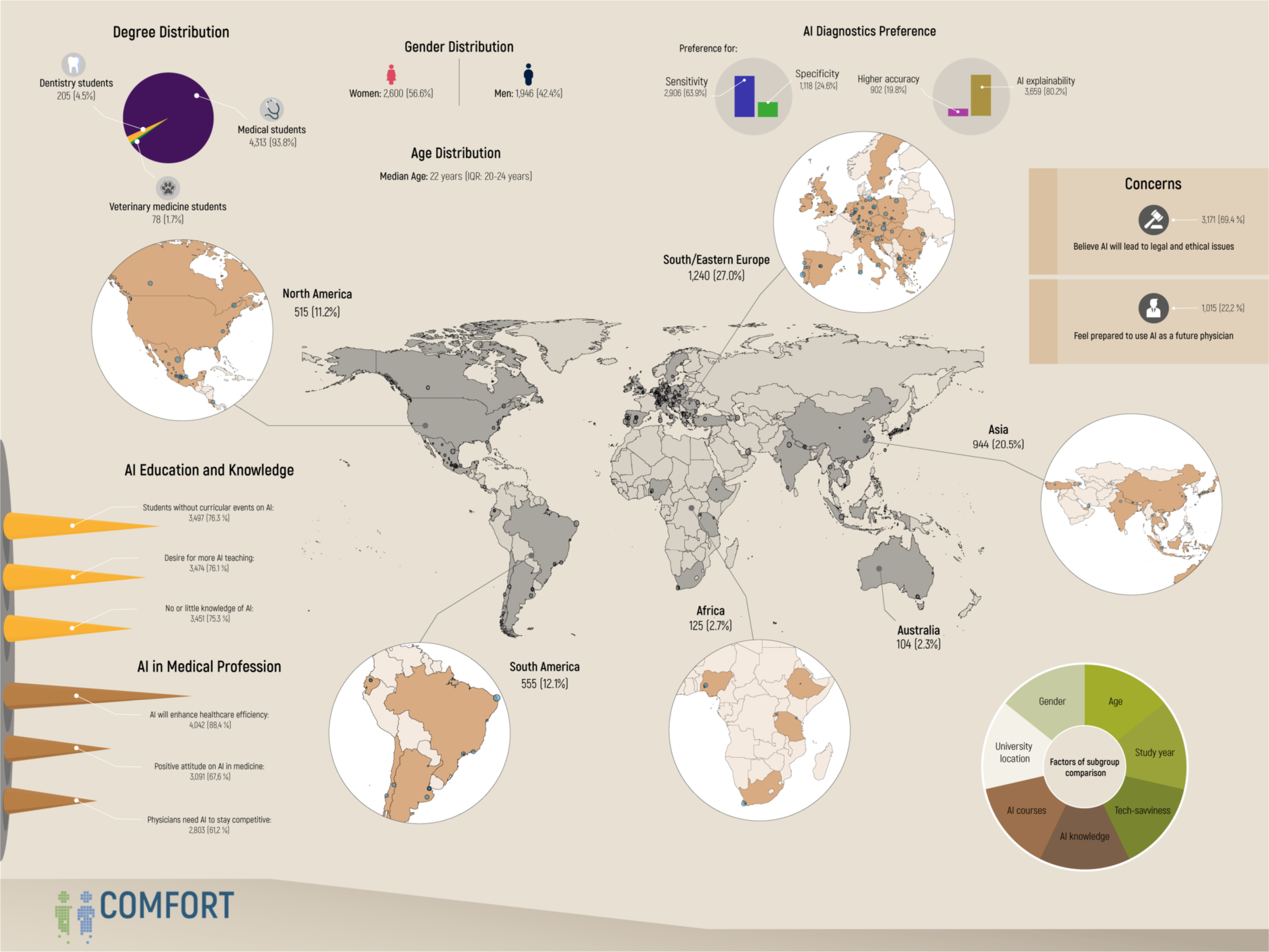

## Introduction

The popularity of artificial intelligence (AI) in healthcare has exponentially risen in recent years, attracting the attention of professionals and students alike [1, 2]. With the debut of OpenAI’s Chat Generative Pre-trained Transformer (ChatGPT) in November 2022, a large language model (LLM)-based chatbot with human-like conversational capabilities, this development has gained further momentum [3, 4]. Although not primarily trained with medical data, ChatGPT-3.5 was barely able to pass the United States Medical Licensing Examination (USMLE), while its successor, GPT-4, surpassed it substantially with a zero short accuracy of 84.3% [5]. Specialized medical LLMs, such as Google’s MedPaLM-2, have advanced the field even further, achieving over 85% accuracy on the MedQA dataset of USMLE-like questions and being the first model to pass the MedMCQA dataset, which includes medical exam questions from the All India Institute for Medical Sciences and the National Eligibility cum Entrance Test [6].

Similarly, specialized machine learning models have demonstrated their potential in clinical applications, achieving expert-level results in areas such as breast cancer screening, pathology detection on chest radiographs, or high accuracy in predicting survival and treatment response [7–10].

Given the range and capabilities of potential applications in medicine, the integration of AI into the medical curriculum holds promise, for example, by supporting the teaching of medical students through explaining medical terminology, answering medical questions, creating intelligent learning plans, providing real-time automated feedback, curating data, or simulating patient conversations [11–14]. In addition, teaching AI in medicine may ensure that medical students are familiar with the AI tools and technologies they will inevitably encounter in their professional lives [15, 16].

On the other hand, the introduction of medical AI into clinical routine and medical education poses new challenges, such as ensuring patient and user autonomy, beneficence, non-maleficence, and justice according to the core biomedical ethical principles [11, 17, 18]. Therefore, especially in a field as critical as medicine, where vital measures can be taught and sensible data is handled, it is essential to include education about both the benefits and risks of AI already in the medical curriculum, preparing students for its qualified and responsible use from their career’s onset [19–21].

While previous studies have partially explored the attitudes of healthcare students towards AI, reporting positive views and a desire for its inclusion in the curriculum, they have been primarily limited by their small sample size, restricted scope, or geographic specificity, precluding a comprehensive international comparison of perceptions [22]. To address these limitations, the present study is an international, multicenter effort to conduct an online survey to assess and compare the perspectives of medical, dental, and veterinary students regarding: 1) their technological literacy and understanding of informatics and AI, 2) the current state of AI in their respective curricula and preferences for AI education, and 3) their attitudes toward the role of AI in their respective fields. In addition, the study seeks to explore regional differences in perspectives.

## Methods

This multicenter cross-sectional study was conducted in accordance with the STROBE statement and received ethical approval from the Institutional Review Board at Charité – University Medicine Berlin (EA4/213/22), serving as the principal institution, in compliance with the Declaration of Helsinki and its later amendments. To ensure participant anonymity, the necessity for informed consent was waived.

### Instrument development and design

Following the Association for Medical Education In Europe (AMEE) Guide, this study aimed to develop an anonymous online survey to assess: 1) the technological literacy and knowledge of informatics and AI, 2) the current state of AI in their respective curricula and preferences for AI education, and 3) the perspectives towards AI in the medical profession among international medicine, dentistry, and veterinary medicine students [23]. To inform instrument development, a literature review of existing publications on the attitudes of medical students towards AI in medicine was independently performed by four reviewers (FB, LH, KKB, LCA), leveraging MEDLINE, Scopus, and Google Scholar databases in December 2022. Studies were selected for review based on the following criteria: 1) the publications were original research articles, 2) the scope aligned with our research objectives and targeted medical students, 3) the survey was conducted in English language, 4) the items were publicly accessible, 5) the measurement of perspectives towards AI was not restricted to a particular medical subfield. Following these criteria, five articles comprising a total of 96 items were identified as relevant to the research scope [24–28]. After a consensus-based discussion, items that did not match our research objectives or overlapped in content were excluded, resulting in 23 remaining items. These items were subsequently tailored to fit the context of medical education and the medical profession.

A review cycle was undertaken with a focus group of medical AI researchers and students, as well as an expert panel including physicians, medical faculty members and educators, AI researchers and developers, and biomedical statisticians (FB, LH, DT, MRM, KKB, LCA, AB, RC, GDV, AH, LJ, AL, PS, LX). The finalized survey consisted of 16 multiple-choice items, eight demographic queries, and one free-field comment section. These items were further refined based on content-based domain samples, and responses were standardized using a four- or five-point Likert scale where applicable.

The preliminary assessment was conducted through cognitive interviews with ten medical students at Charité – University Medicine Berlin to evaluate the scale’s comprehensiveness and overall length. The feedback resulted in two rewordings and one item removal, finalizing the survey with 15 multiple-choice items and eight demographic queries supported by one free-field comment section.

Using REDCap (Research Electronic Data Capture) hosted at Charité – University Medicine Berlin, the English survey was subsequently disseminated through the medical student newsletter at Charité and deactivated after receiving responses from 50 medical students who served as the pilot study group and were not included in the final participant pool [29, 30]. After psychometric validation, participating sites distributed the REDCap online survey among medical, dental, and veterinary students at their faculty. Due to the large number of Spanish-speaking sites, a separate Spanish online version of the survey was employed using paired forward and backward translation with reconciliation by two bilingual medical professionals (LG, JSPO). Depending on their faculty location, participating sites distributed either the English or Spanish online survey via their faculty newsletters and courses using a QR code or the direct website link. The survey was available for participation from April to October 2023.

### Inclusion and exclusion criteria

Inclusion criteria consisted of students at least 18 years of age, actively enrolled in a (human) medicine, dentistry, or veterinary medicine degree program, who responded to the survey during its open period and were proficient in either English or Spanish, depending on their faculty location. Students from unrelated degree programs, postgraduates, respondents who did not indicate information about their course, and respondents who started the survey but did not answer any multiple-choice items were excluded from the analysis. Partial missing responses to survey items resulted in exclusion from each subanalysis.

### Statistical analysis

Statistical analyses were performed with SPSS Statistics 25 (version 28.0.1.0) and R (version 4.2.1), using the “tidyverse”, “rnaturalearth”, and “sf” packages [31–34]. The Kolmogorov-Smirnov test was used to test for normal distribution. Categorical and ordinal data were reported as frequencies with percentages. Medians and interquartile ranges (IQR) were reported for non-parametric continuous data. Variances were reported for items in Likert scale format. The response rate was derived from the overall student enrollment numbers at each faculty according to the faculty websites or the Times Higher Education World University Rankings 2024 due to the unavailability of official data on enrolled medical, dentistry, or veterinary students. In the pilot study group, item reliability was measured using Cronbach’s alpha, with values above 0.7 interpreted as acceptable internal consistency.

Explanatory factor analysis was used to examine the structure and subscales of the instrument, using an eigenvalue cutoff of 1 for item extraction. Items with factor loadings of 0.4 or higher were retained. Data suitability for structural evaluation was assessed using the Kaiser-Meyer-Olkin measure and Bartlett’s test of sphericity. For geographical subgroup analysis, respondents were categorized based on their faculty location (Global North versus Global South) according to the United Nations’ Finance Center for South-South Cooperation [35].

Additionally, participants were grouped into continents based on the United Nations geoscheme [36]. Due to the substantial number of European participants, students in North/West and South/East Europe were analyzed separately. Further subgroup analyses based on gender, age, academic year, technological literacy, self-reported AI knowledge, and previous curricular AI events can be found in the supporting information. The Mann-Whitney U-test was employed for subgroup analyses of two independent non-parametric samples. For continental comparison, the Kruskal-Wallis one-way analysis of variance and Dunn-Bonferroni post hoc test were performed. To estimate effect size, we calculated r, with 0.5 indicating a large effect, 0.3 a medium effect, and 0.1 a small effect [37]. An asymptotic two-sided p-value below 0.05 was considered statistically significant.

## Results

### Pilot study

The median age of the pilot study group was 24 years (IQR: 21-26 years). 58% of participants identified as female (N=29), 38% as male (N=19), and 4% (N=2) did not report their gender. The median current academic year was 2 (IQR: 2-4 years) out of 6 total academic years. Internal consistency for our scale’s dimensions ranged from acceptable to good, as indicated by Cronbach’s alpha. The section on “Technological literacy and knowledge of informatics and AI” registered an alpha of 0.718, while the section “Current state of AI in the curriculum and preferences for AI education” scored an alpha of 0.726, both displaying acceptable internal consistency. A Cronbach’s alpha value of 0.825 for the “Perspectives towards AI in the medical profession” section denoted good internal consistency. The Kaiser-Meyer-Olkin measure for sampling adequacy was 0.801, confirming the sample’s representational validity. Bartlett’s test of sphericity returned a p-value of less than 0.001, validating the chosen method for factor analysis. Factor analysis yielded a structure comprising 15 items across three dimensions, collectively explaining 54% of the total variance. Factor loadings for individual items ranged from 0.495 for “Which of these technical devices do you use at least once a week?” to 0.888 for “What is your general attitude toward the application of artificial intelligence (AI) in medicine?”.

### Study cohort

Between the first of April and the first of October 2023, 4,900 responses were recorded, of which 4,345 (88.7%) were collected via the English survey and 555 (11.3%) via the Spanish survey version. Of these, 283 (5.8%) respondents reported degrees other than medicine, dentistry, or veterinary medicine or indicated that they had completed their studies, while 21 (0.4%) did not respond to any multiple-choice item or did not indicate their degree. The final study cohort comprised 4,596 participants from 192 faculty and 48 countries, of whom 4,313 (93.8%) were medical, 205 (4.5%) dentistry, and 78 (1.7%) veterinary medicine students. Of 5,575,307 enrolled students from all degrees at the 183 (95.3%) participating faculties in which the total enrollment number was publicly available, the survey achieved an average response rate of 0.2% (standard deviation: 0.4%). Most respondents studied in Southern/Eastern European (N=1,240, 27.0%) countries, followed by Northern/Western Europe (N=1,110, 24.2%), Asia (N=944, 20.5%), South America (N=555, 12.1%), North America (N=515, 11.2%), Africa (N=125, 2.7%), and Australia (N=104, 2.3%). Please refer to Fig 1 to view the distribution of participating institutions in relation to the number of participants on a world map. A detailed list of survey participants divided by country, faculty, city, degree, number of enrolled students, and response rate is provided in the supplemental material (S1 Table). The median age of the study population was 22 years (IQR: 20-24 years). 56.6% of the participants were female (N=2,600) and 42.4% male (N=1,946), with a median academic year of 3 (IQR: 2-5 years). Full descriptive data, including items on technological literacy and preferences for AI teaching in the medical curriculum, are displayed in Table 1.

**Fig 1.**
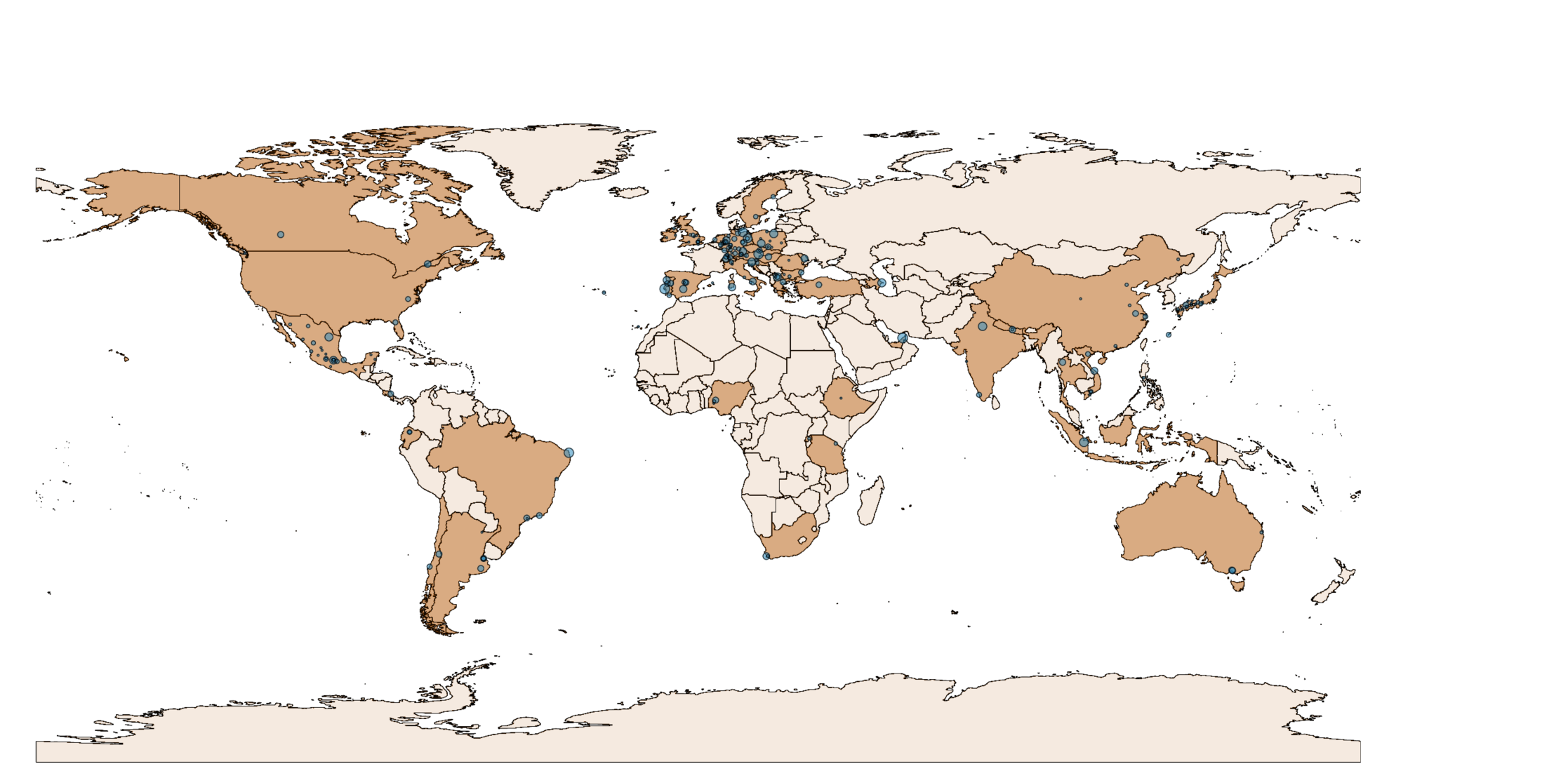
The world map displays the geographical distribution of participating institutions (blue dots) in relation to the number of respondents per institution.

Any free field comments of the survey participants are listed in the supplements (S2 Table), with selected comments highlighted in Fig 2.

**Fig 2.**
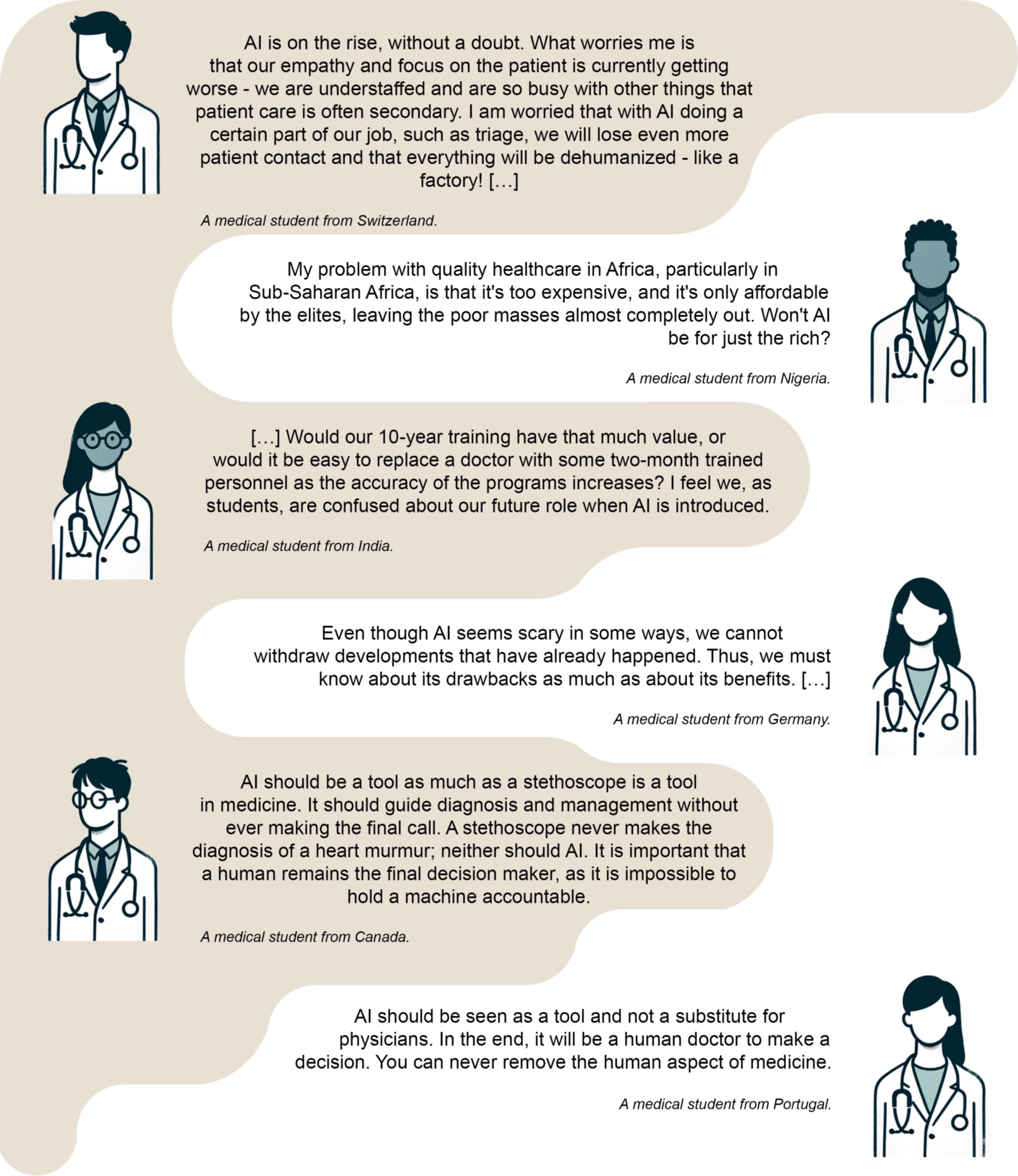
Diverse perspectives from medical students on the integration of artificial intelligence (AI) in healthcare education and practice. The selected quotes reflect a range of sentiments, from concerns about dehumanization and potential challenges in low-resource settings to viewing AI as a beneficial tool that complements rather than replaces the human touch in medicine.

**Table 1.** Descriptive data of the study population and results of the questions about tech- savviness and topic preferences for AI teaching in the medical curriculum.

| Item | Value |
| --- | --- |
| Gender (N=4,594) | N (%) |
| Female | 2,600 (56.6) |
| Male | 1,946 (42.4) |
| Diverse | 25 (0.5) |
| Prefer not to disclose | 23 (0.5) |
| Age (N=4,571) | Years, median (IQR) |
|  | 22 (20-24) |
| Current academic year (N=4,473) | Years, median (IQR) |
|  | 3 (2-5) |
| Total academic years (N=4,315) | Years, median (IQR) |
|  | 6 (6-6) |
| Which of these technical devices do you use at least once a week? (N=4,596) | N (%) |
| Smartphone | 4,406 (95.9) |
| PC/laptop | 4,020 (87.5) |
| Game console (e.g., PlayStation, Switch) | 511 (11.1) |
| Tablet (e.g., iPad) | 2,172 (47.3) |
| E-reader | 325 (7.1) |
| Smartwatch | 1,033 (22.5) |
| None | 9 (0.0) |
| Have you already programmed code? (N=4,585<sup>a</sup>) | N (%) |
| Yes | 912 (19.9) |
| C | 85 (1.9) |
| C++ | 155 (3.4) |
| C# | 37 (0.8) |
| CSS | 32 (0.7) |
| HTML | 107 (2.3) |
| Java | 166 (3.6) |
| JavaScript | 91 (2.0) |
| MATLAB | 35 (0.8) |
| Pascal | 17 (0.4) |
| PHP | 24 (0.5) |
| Python | 382 (8.3) |
| R | 284 (6.2) |
| SQL | 11 (0.2) |
| Visual Basic | 14 (0.3) |
| No | 3,673 (80.1) |
| What would you like to learn about artificial intelligence (AI) as part of your medical curriculum? (N=4,596) | N (%) |
| Theory and background (e.g., mathematical basics) | 1,549 (33.7) |
| Practical skills (e.g., learning programming languages; solving medical problems with AI) | 3,515 (76.5) |
| History and development | 827 (18.0) |
| Legal and ethical aspects | 2,518 (54.8) |
| Future perspectives of AI in medicine | 3,278 (71.3) |
| No preference | 162 (3.5) |
| None | 12 (0.3) |

<sup>a</sup> To enhance data presentation, programming languages with a sample size of fewer than 10
respondents were omitted.
IQR, interquartile range.

### Collective perceptions towards artificial intelligence

Table 2 displays the survey results for Likert scale items. Students generally reported a rather or extremely positive attitude towards the application of AI in medicine (3,091, 67.6%). The highest positive attitude towards AI in the medical profession was recorded for the item “How do you estimate the effect of artificial intelligence (AI) on the efficiency of healthcare processes in the next 10 years?” with 4,042 respondents (88.4%) estimating a moderate or great improvement. Contrarily, 3,171 students (69.4%) rather or completely agreed with the item “The use of artificial intelligence (AI) in medicine will increasingly lead to legal and ethical conflicts.”. Regarding AI education and knowledge, 3,451 students (75.3%) reported no or little knowledge of AI, and 3,474 (76.1%) rather or completely agreed that they would like to have more teaching on AI in medicine as part of their curricula. On the other hand, 3,497 (76.3%) students responded that they did not have any curricular events on AI as part of their degree, as illustrated on the country level in Fig 3. Variability in responses was observed, ranging from 0.279 for the item “How would you rate your general knowledge of artificial intelligence (AI)?” —measured on a four-point Likert scale— to 1.372 for “With my current knowledge, I feel sufficiently prepared to work with artificial intelligence (AI) in my future profession as a physician.”. Notably, the items capturing the trade-offs in medical AI diagnostics revealed that most students preferred AI explainability (N=3,659, 80.2%) over a higher accuracy (N=902, 19.8%) and higher sensitivity (N=2,906, 63.9%) over higher specificity (N=1,118, 24.6%) or equal sensitivity/specificity (N=524, 11.5%), as visualized in Fig 4.

**Table 2.** Survey results of Likert scale format items on attitudes towards the medical degree, AI in the medical profession, AI education and knowledge, and results of the regional comparisons.

| Item | Total responses |  |  |  |  |  | Regional comparison Global North/Global South |  |  |  |
| --- | --- | --- | --- | --- | --- | --- | --- | --- | --- | --- |
|  | Completely disagree/<br>Extremely negative/<br>Great deterioration<br>N (%) | Rather disagree/<br>Rather negative/<br>Moderate deterioration<br>N (%) | Neutral/No effect<br>N (%) | Rather agree/<br>Rather positive/<br>Moderate improvement<br>N (%) | Completely agree/<br>Extremely positive/<br>Great improvement<br>N (%) | Variance | Global North<br>Median (IQR) | Global South<br>Median (IQR) | P-value<sup>a</sup> | r |
| **Attitude towards medical studies** | - | - | - | - | - | - | - | - | - | - |
| What is your current general attitude toward your medical studies? | 48 (1.0) | 216 (4.7) | 899 (19.6) | 2,597 (56.7) | 819 (17.9) | 0.638 | N=3,162<br>4 (4-4) | N=1,417<br>4 (3-4) | <.001 | .095 |
| **Perspectives towards AI in the medical profession** | - | - | - | - | - | - | - | - | - | - |
| What is your general attitude toward the application of artificial intelligence (AI) in medicine? | 60 (1.3) | 307 (6.7) | 1,118 (24.4) | 2,286 (50.0) | 805 (17.6) | 0.748 | N=3,164<br>4 (3-4) | N=1,412<br>4 (3-4) | .707 | .006 |
| How do you estimate the effect of artificial intelligence (AI) on the efficiency of healthcare processes in the next 10 years? | 48 (1.1) | 194 (4.2) | 286 (6.3) | 2,687 (58.8) | 1,355 (29.6) | 0.610 | N=3,155<br>4 (4-5) | N=1,415<br>4 (4-5) | <.001 | .061 |
| The use of artificial intelligence (AI) in medicine will increasingly lead to legal and ethical conflicts. | 73 (1.6) | 370 (8.1) | 961 (21.0) | 1,869 (40.9) | 1,302 (28.5) | 0.944 | N=3,159<br>4 (3-5) | N=1,416<br>4 (3-4) | <.001 | .185 |
| What is your view on the influence of artificial intelligence (AI) on the profession of physicians? AI will affect the everyday life of physicians in a way that is... | 50 (1.1) | 323 (7.1) | 1,027 (22.5) | 2,676 (58.5) | 495 (10.8) | 0.630 | N=3,157<br>4 (3-4) | N=1,414<br>4 (3-4) | .002 | .046 |
| How would you rate artificial intelligence (AI) software being available to physicians as a second opinion on medical issues? | 89 (1.9) | 428 (9.4) | 1,026 (22.5) | 2,281 (50.0) | 741 (16.2) | 0.842 | N=3,155<br>4 (3-4) | N=1,410<br>4 (3-4) | .019 | .035 |
| I think working with artificial intelligence (AI) as a physician is necessary to stay competitive. | 135 (3.0) | 523 (11.4) | 1,115 (24.4) | 1,988 (43.4) | 815 (17.8) | 0.998 | N=3,160<br>4 (3-4) | N=1,416<br>4 (3-4) | <.001 | .049 |
| With my current knowledge, I feel sufficiently prepared to work with artificial intelligence (AI) in my future profession as a physician. | 1,003 (21.9) | 1,649 (36.0) | 910 (19.9) | 740 (16.2) | 275 (6.0) | 1.372 | N=3,161<br>2 (1-3) | N=1,416<br>3 (2-4) | <.001 | .162 |
| **AI education and knowledge level** | - | - | - | - | - | - | - | - | - | - |
| I would like to have more teaching on artificial intelligence (AI) in medicine as part of my studies. (N=4,565) | 86 (1.9) | 191 (4.2) | 814 (17.8) | 2,033 (44.5) | 1,441 (31.6) | 0.831 | N=3,155<br>4 (4-5) | N=1,410<br>4 (3-5) | .107 | .024 |
| As part of my studies, there are curricular events on artificial intelligence (AI) in medicine. (N=4,581) | **No**<br>**N (%)** | **Yes; 1-5 hours in total**<br>**N (%)** | **Yes; >5-10 hours in total**<br>**N (%)** | **Yes; >10-20 hours in total**<br>**N (%)** | **Yes; >20 hours in total**<br>**N (%)** | - | - | - | - | - |
|  | 3,497 (76.3) | 820 (17.9) | 178 (3.9) | 48 (1.0) | 38 (0.8) | 0.458 | N=3,165<br>1 (1-1) | N=1,416<br>1 (1-2) | <.001 | .090 |
| How would you rate your general knowledge of artificial intelligence (AI)? (N=4,585) | **No knowledge (never heard of AI)**<br>**N (%)** | **Little knowledge (e.g., documentary seen on TV)**<br>**N (%)** | **Good knowledge (e.g., read several journal articles on AI)**<br>**N (%)** | **Expert (e.g., involved in AI research/development)**<br>**N (%)** | - | - | - | - | - | - |
|  | 170 (3.7) | 3,281 (71.6) | 1,064 (23.2) | 70 (1.5) | - | 0.279 | N=3,167<br>2 (2-3) | N=1,418<br>2 (2-2) | <.001 | .025 |

**Table 2. (Continued)**
| Item | Regional comparison continental |  |  |  |  |  |  |  | Significant differences in Dunn-Bonferroni post hoc analysis<br>1, North/Western Europe; 2, South/Eastern Europe; 3, Asia; 4, North America; 5, South America; 6, Africa; 7, Australia |
| --- | --- | --- | --- | --- | --- | --- | --- | --- | --- |
|  | North/Western Europe<br>Median (IQR) | South/Eastern Europe<br>Median (IQR) | Asia<br>Median (IQR) | North America<br>Median (IQR) | South America<br>Median (IQR) | Africa<br>Median (IQR) | Australia<br>Median (IQR) | P-value<sup>b</sup> |  |
| **Attitude towards medical studies** | - | - | - | - | - | - | - | - | - |
| What is your current general attitude toward your medical studies? | N=1,108<br>4 (4-4) | N=1,230<br>4 (4-4) | N=941<br>4 (3-4) | N=514<br>4 (4-4) | N=555<br>4 (3-4) | N=124<br>4 (4-5) | N=104<br>4 (4-4) | <.001 | 1-4 (p=.034; r=.078); 2-3 (p=.021; r=.071); 3-4 (p<.001; r=.132); 3-6 (p=.014; r=.104); 4-5 (p=.002; r=.121) |
| **Perspectives towards AI in the medical profession** | - | - | - | - | - | - | - | - | - |
| What is your general attitude toward the application of artificial intelligence (AI) in medicine? | N=1,109<br>4 (3-4) | N=1,231<br>4 (3-4) | N=937<br>4 (3-4) | N=514<br>4 (3-4) | N=554<br>4 (4-4) | N=124<br>4 (3-4) | N=104<br>4 (3-4) | <.001 | 1-5 (p=.004; r=.091); 1-7 (p=.038; r=.090); 2-5 (p<.001; r=.104); 3-5 (p<.001; r=.124); 4-7 (p=.026; r=.130); 5-7 (p<.001; r=.187) |
| How do you estimate the effect of artificial intelligence (AI) on the efficiency of healthcare processes in the next 10 years? | N=1,107<br>4 (4-5) | N=1,223<br>4 (4-5) | N=939<br>4 (4-5) | N=515<br>4 (4-5) | N=555<br>4 (4-5) | N=124<br>4 (4-5) | N=104<br>4 (4-4) | .001 | 1-3 (p=.071); 1-4 (p=.012; r=.085) |
| The use of artificial intelligence (AI) in medicine will increasingly lead to legal and ethical conflicts. | N=1,108<br>4 (4-5) | N=1,227<br>4 (4-5) | N=941<br>4 (3-4) | N=515<br>4 (3-4) | N=554<br>4 (3-4) | N=124<br>4 (3-5) | N=103<br>4 (4-5) | <.001 | 1-3 (p<.001; r=.216); 1-4 (p<.001; r=.202); 1-5 (p<.001; r=.301); 2-3 (p<.001; r=.175); 2-4 (p<.001; r=.161); 2-5 (p<.001; r=.258); 3-5 (p=.003; r=.099); 3-7 (p<.001; r=.194); 4-5 (p=.018; r=.102); 4-7 (p<.001; r=.243); 5-6 (p<.001; r=.174); 5-7 (p<.001; r=.311); 6-7 (p=.048; r=.203) |
| What is your view on the influence of artificial intelligence (AI) on the profession of physicians? AI will affect the everyday life of physicians in a way that is... | N=1,109<br>4 (3-4) | N=1,225<br>4 (3-4) | N=940<br>4 (3-4) | N=515<br>4 (3-4) | N=553<br>4 (4-4) | N=124<br>4 (3-4) | N=104<br>4 (3-4) | <.001 | 1-3 (p<.001; r=.106); 2-5 (p<.001; r=.128); 3-4 (p<.001; r=.127); 3-5 (p<.001; r=.164); 5-7 (p=.012; r=.135) |
| How would you rate artificial intelligence (AI) software being available to physicians as a second opinion on medical issues? | N=1,108<br>4 (3-4) | N=1,226<br>4 (3-4) | N=936<br>4 (3-4) | N=513<br>4 (3-4) | N=553<br>4 (4-4) | N=124<br>4 (3-4) | N=103<br>4 (3-4) | <.001 | 1-2 (p=.008; r=.073); 1-3 (p<.001; r=.125); 1-6 (p=.020; r=.094); 1-7 (p=.017; r=.096); 2-5 (p<.001; r=.128); 3-4 (p=.034; r=.083); 3-5 (p<.001; r=.185); 4-5 (p=.014; r=.104); 5-6 (p<.001; r=.172); 5-7 (p<.001; r=.173) |
| I think working with artificial intelligence (AI) as a physician is necessary to stay competitive. | N=1,108<br>4 (3-4) | N=1,228<br>4 (3-4) | N=940<br>4 (3-4) | N=514<br>4 (3-4) | N=555<br>4 (4-4) | N=124<br>4 (3-4) | N=104<br>4 (3-4) | <.001 | 1-5 (p<.001; r=.124); 2-5 (p<.001; r=.172); 3-5 (p<.001; r=.124); 4-5 (p<.001; r=.146); 5-7 (p=.027; r=.126) |
| With my current knowledge, I feel sufficiently prepared to work with artificial intelligence (AI) in my future profession as a physician. | N=1,108<br>2 (1-2) | N=1,228<br>2 (2-3) | N=940<br>3 (2-4) | N=515<br>2 (2-4) | N=555<br>2 (2-3) | N=124<br>3 (2-4) | N=104<br>2 (2-3) | <.001 | 1-2 (p<.001; r=.187); 1-3 (p<.001; r=.531); 1-4 (p<.001; r=.259); 1-5 (p<.001; r=.114); 1-6 (p<.001; r=.197); 2-3 (p<.001; r=.342); 2-4 (p=.011; r=.083); 3-4 (p<.001; r=.243); 3-5 (p<.001; r=.398); 3-6 (p<.001; r=.132); 3-7 (p<.001; r=.248); 4-5 (p<.001; r=.158); 5-6 (p=.001; r=.159); 6-7 (p=.037; r=.208) |
| **AI education and knowledge level** | - | - | - | - | - | - | - | - | - |
| I would like to have more teaching on artificial intelligence (AI) in medicine as part of my studies. | N=1,106<br>4 (4-5) | N=1,226<br>4 (3-5) | N=933<br>4 (3-4) | N=514<br>4 (4-5) | N=555<br>4 (4-5) | N=124<br>4 (3-5) | N=104<br>4 (3-5) | <.001 | 1-2 (p=.003; r=.078); 1-3 (p<.001; r=.144); 2-3 (p=.049; r=.066); 2-4 (p<.001; r=.091); 2-5 (p<.001; r=.122); 3-4 (p<.001; r=.174); 3-5 (p<.001; r=.210) |
| As part of my studies, there are curricular events on artificial intelligence (AI) in medicine. | - | - | - | - | - | - | - | - | - |
|  | N=1,109<br>1 (1-1) | N=1,233<br>1 (1-1) | N=941<br>1 (1-2) | N=513<br>1 (1-1) | N=554<br>1 (1-1) | N=124<br>1 (1-1) | N=104<br>1 (1-1) | <.001 | 1-3 (p<.001; r=.172); 2-3 (p<.001; r=.202); 3-4 (p<.001; r=.141); 3-5 (p<.001; r=.225); 3-6 (p<.001; r=.154); 3-7 (p=.017; r=.103) |
| How would you rate your general knowledge of artificial intelligence (AI)? | - | - | - | - | - | - | - | - | - |
|  | N=1,109<br>2 (2-2) | N=1,233<br>2 (2-3) | N=942<br>2 (2-3) | N=515<br>2 (2-3) | N=555<br>2 (2-2) | N=124<br>2 (2-3) | N=104<br>2 (2-3) | <.001 | 1-4 (p=.015; r=.089) |

<sup>a</sup> Compares student responses in Global North and South using the Mann-Whitney U-test.
<sup>b</sup> Compares student responses across all regions using the Kruskal-Wallis one-way analysis of variance.
IQR, interquartile range.

**Fig 3.**
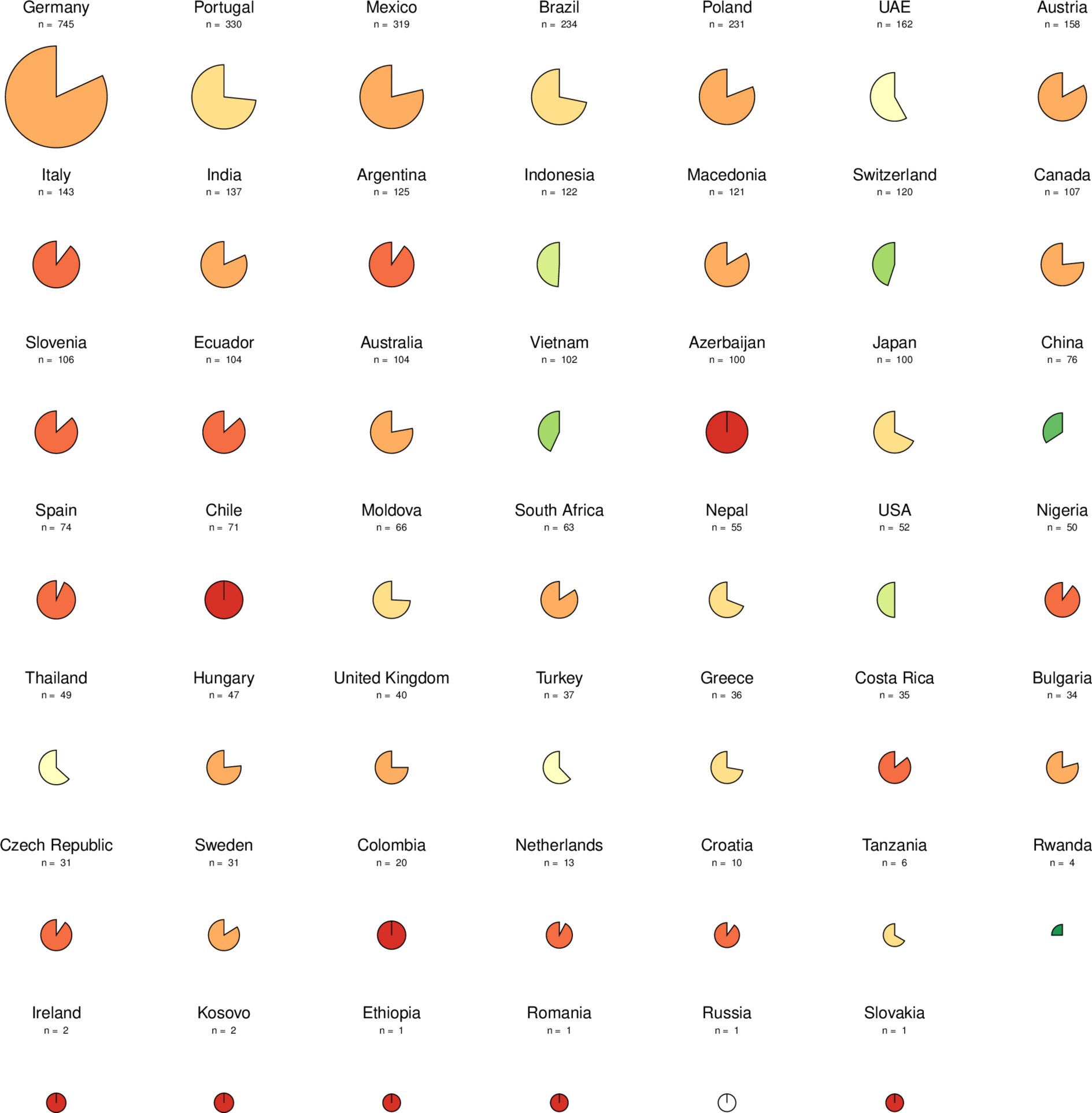
Pie charts illustrating student responses at the country level for the item “As part of my studies, there are curricular events on artificial intelligence (AI) in medicine.”. A more filled, darker red chart indicates a higher proportion of students reporting no AI events, while a less filled, greener chart indicates fewer students reporting the absence of AI events. The missing portion of each chart displays the proportion of students who reported AI events, regardless of the duration. An all-white pie chart indicates that all students reported AI events in the medical curriculum. The absolute number of responses per country is shown above each chart. Analysis of the pie charts from countries with a representative sample of at least 50 respondents reveals that, among 28 nations, only four (Indonesia, Switzerland, Vietnam, and China) exhibited over 50% of students reporting the inclusion of AI events within their medical curriculum. Data from the USA displayed an equal proportion of students reporting the presence or absence of AI events in their curriculum (50% each). The residual 23 countries, encompassing Germany, Portugal, Mexico, Brazil, Poland, UAE, Austria, Italy, India, Argentina, Macedonia, Canada, Slovenia, Ecuador, Australia, Azerbaijan, Japan, Spain, Chile, Moldova, South Africa, Nepal, and Nigeria, had a lower proportion of students reporting the integration of AI in the medical curriculum. UAE, United Arab Emirates; USA, United States of America.

**Fig 4.**
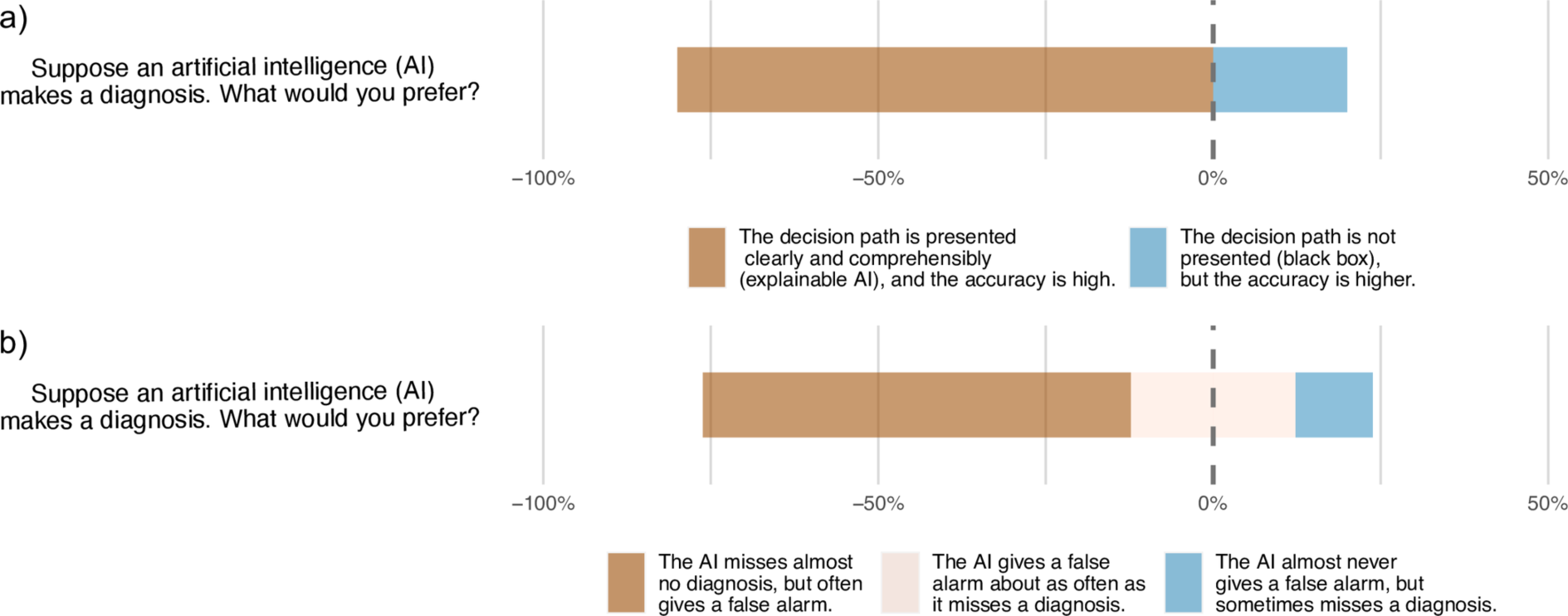
Gantt diagrams depicting medical students’ preferences in AI diagnostics. a) AI explainability (N=3,659, 80.2%) versus higher accuracy (N=902, 19.8%) and b) higher sensitivity (N=2,906, 63.9%) versus higher specificity (N=1,118, 24.6%) or equal sensitivity and specificity (N=524, 11.5%).

### Regional comparisons

Please refer to Table 2 to view the results of the comparison between responses from the Global North and South and at the continental level. Perceptions between the Global North and South differed significantly for nine Likert scale format items. The highest effect size was observed for the item on AI increasing ethical and legal conflicts, with respondents from the Global North indicating a higher agreement (median: 4, IQR: 3-5) compared to those from the Global South (median: 4, IQR: 3-4; r=0.185). Notably, Global South students felt more prepared to use AI in their future practice (median: 3, IQR: 2-4) compared to their Global North counterparts (median: 2, IQR: 1-3; r=0.162) and reported longer AI-related curricular events (median: 1, IQR: 1-2; Global North: median: 1, IQR: 1-1; r=0.090).

Conversely, Global North students rated their AI knowledge higher (median: 2, IQR: 2-3; Global South: median: 2, IQR: 2-2; r=0.025).

For continental comparison, the Kruskal-Wallis one-way analysis of variance revealed significantly different Likert scale responses across all survey items. Subsequent Dunn-Bonferroni post hoc analysis displayed various significant differences in Likert scale responses for pairwise regional comparisons, while median and IQR remained largely consistent. Considering only medium to large effect sizes, the item “The use of artificial intelligence (AI) in medicine will increasingly lead to legal and ethical conflicts.” yielded an r of 0.301 when comparing Northern/Western European (median: 4, IQR: 4-5) and South American participants (median: 4, IQR: 3-4), and an r of 0.311 between South American and Australian participants (median: 4, IQR: 4-5). Similarly, the statement “With my current knowledge, I feel sufficiently prepared to work with artificial intelligence (AI) in my future profession as a physician.” displayed strong effect sizes in comparisons between North/West Europe (median: 2, IQR: 1-2) and Asia (median: 3, IQR: 2-4; r=0.531), South/East Europe (median: 2, IQR: 2-3) and Asia (r=0.342), and South America (median: 2, IQR: 2-3) and Asia (r=0.398).

## Discussion

This multicenter survey of 4,596 medical, dental, and veterinary students from 192 faculties in 48 countries reveals an optimistic outlook of medical students about the future role of AI in healthcare practice. However, this optimism is mixed with concerns, with a majority foreseeing increasing ethical and legal challenges as AI technologies become more integrated into medical practice and feeling inadequately prepared to use AI in their later careers. In terms of AI knowledge and education, our results demonstrate that most students report no or little AI knowledge and favor the inclusion of AI teaching in their curriculum, while over three-quarters did not have any events on AI as part of their studies. The subgroup analyses revealed regional differences in perceptions, although predominantly with small effect sizes.

To date, multiple studies have examined healthcare students’ attitudes towards AI in medicine [22]. Focusing on studies with large sample sizes of at least 500 medical or dental students, predominantly positive attitudes toward the application of AI in medicine were reported [24, 38–42]. For instance, a 2021 study by Bisdas et al., spanning 2,495 medical and 638 dental students in 63 countries, most of whom studying in Libya (N=790), Jordan (N=450), and the United Kingdom (N=331), found that 88.2% (N=2,766) of respondents either strongly or somewhat agree that AI will generally enhance medical practice, while about half (N=1,655, 52.8%) indicated that they would usually or always incorporate AI in their future practice [24]. Similarly, a 2022 survey of 2,981 medical students in Turkey by Civaner et al. demonstrated that 85.8% (N=2,558) of participants acknowledge AI’s role in facilitating physicians’ access to information, while 74.4% (N=2,218) of students affirm that widespread AI usage would make them better physicians and 70.5% (N=2,102) believe that AI application would lead to reduced errors [38]. Another large-scale survey conducted in 2020 among 1,103 dental students in Turkey by Yüzbaşıoğlu et al. demonstrated a significant agreement (N=945, 85.7%) on the prospective major advancements in dentistry and medicine through AI, which was similar to a 2022 study by Swed et al., comprising a cohort of 1,251 medical students and 243 physicians in Syria, with 1,317 respondents (87.4%) either strongly agreeing or agreeing on the crucial role of AI in the medical field [39, 40]. These findings are consistent with those of our study, in which most participants generally expressed positive attitudes toward the use of AI in medicine, the increase in efficiency of healthcare processes through AI, the positive impact of AI on the medical profession, and the availability of AI as a second opinion on medical issues.

Despite these positive attitudes, our study revealed a pronounced deficit in both self-reported AI knowledge and the integration of AI education into the medical curriculum, with about three-quarters of participants reporting little or no AI knowledge, no curricular events on AI, and the desire for AI teaching, respectively. However, the lack of AI knowledge and education among medical and dentistry students is not a novel phenomenon. Depending on the study and item design, self-reported AI knowledge in the literature ranges from 2.8% of 2,981 medical students in Turkey in 2022 who reported feeling informed about the use of AI in medicine to 51.8% of 900 medical students in Jordan in 2021 who indicated having read articles about AI or machine learning in the past two years [24, 38–42]. On the other hand, the reported prevalence of AI training in the medical curriculum ranges, for instance, from 9.2% in a 2020 survey of 484 medical students in the United Kingdom up to 24.4% in a 2022 study among 2,981 medical students in Turkey, although variations in item designs and demographic contexts hinder a comprehensive longitudinal analysis [25, 38, 40, 41, 43]. In our study, less than 18% (N=5) of countries with a sample size of 50 or more participants had a higher or equal proportion of students reporting any duration of AI teaching, pointing to a persistent deficit in medical AI education across various demographic landscapes.

Consequently, the incorporation of AI into medical education on a broader national or international scale is limited, and the adoption of frameworks, certification programs, interdisciplinary collaborations, modules, and formal lectures seems still to be at an early stage [18, 44–46].

In terms of educational preferences, most of the participants in our study indicated their interest in learning practical skills, followed by future perspectives and legal and ethical aspects of medical AI. On the other hand, the majority rather or completely agreed with the item that AI will increasingly lead to legal and ethical conflicts in the medical profession.

Similarly, a 2021 survey by Mehta et al. of 321 medical students at four medical schools in Canada found that 95% of medical students surveyed believe AI would introduce new ethical and social challenges, whereas Civaner et al. reported in 2022 that 93.8% of 2,981 medical students in Turkey consider AI training to address the ethical dilemmas raised by AI a necessary component of their education [38, 47]. This underscores the great potential of AI education to not only improve medical students’ oversight, knowledge, and practical skills in using AI but also to educate about ethical, legal, and societal implications — topics that are also addressed in other AI education frameworks, such as the United Nations Educational, Scientific and Cultural Organization K-12 AI curricula report [48].

In our subgroup analysis of respondents across continents, two items displayed moderate to large effect sizes. First, participants from South America were less likely to agree that the use of medical AI will increase ethical and legal conflicts compared to participants from Northern/Western Europe and Australia. Yet, students’ median responses in these regions were identical. Thus, the level of effect size primarily reflects outliers rather than a uniform regional disparity in opinion. Second, Asian students reported being better prepared to work with AI in their future careers. Although these differences in perceived preparedness could be driven by different national AI policies and educational strategies as well as macroeconomic factors, our study design and varying sample sizes across regions complicate a causal analysis [49, 50]. However, it is noteworthy that three of four countries in which more than 50% of medical students reported receiving AI training were located on the Asian continent, which may have contributed to Asian students indicating a higher preparedness towards applying AI in their future profession.

Finally, we observed a strong preference among medical students for AI systems that are explainable rather than highly accurate. This mirrors the growing emphasis on ’Explainable AI’ in the medical field and underscores the urgent need for developing AI algorithms that transparently disclose their decisions and limitations, especially when applied to the medical domain, to promote trust and acceptance among healthcare students, professionals, and patients [51–54].

This study has limitations. First, the uneven regional distribution of participants potentially biased results in favor of overrepresented regions. Additionally, the online design and language availability in either English or Spanish may have introduced selection bias, precluding students without internet access, as well as students who are not proficient in either language. Another potential source of selection bias could be that respondents with a specific interest in or experience with AI were more likely to participate in the survey.

Furthermore, the calculated response rate appeared to be rather low due to the lack of data on the number of students enrolled in each medical discipline for most participating institutions. Consequently, we derived the response rate using the total student enrollment numbers, which significantly underestimated the true rate of participation among medical students as it assumes that all students within each institute received an invitation to participate. Moreover, the presence of 20 institutions with fewer than 50 student respondents has skewed the response rate further downward.

In conclusion, our study —the currently largest survey of medical students’ perceptions towards AI in healthcare education and practice—reveals a broadly optimistic view of AI’s role in healthcare. It draws on insights from students with diverse geographical, sociodemographic, and cultural backgrounds, underlining the critical need for AI education in medical curricula around the world and identifying a universal challenge and opportunity: to adeptly prepare healthcare students for a future that integrates AI into healthcare practice.

## Acknowledgments

Members of the COMFORT consortium (alphabetically listed by surname):

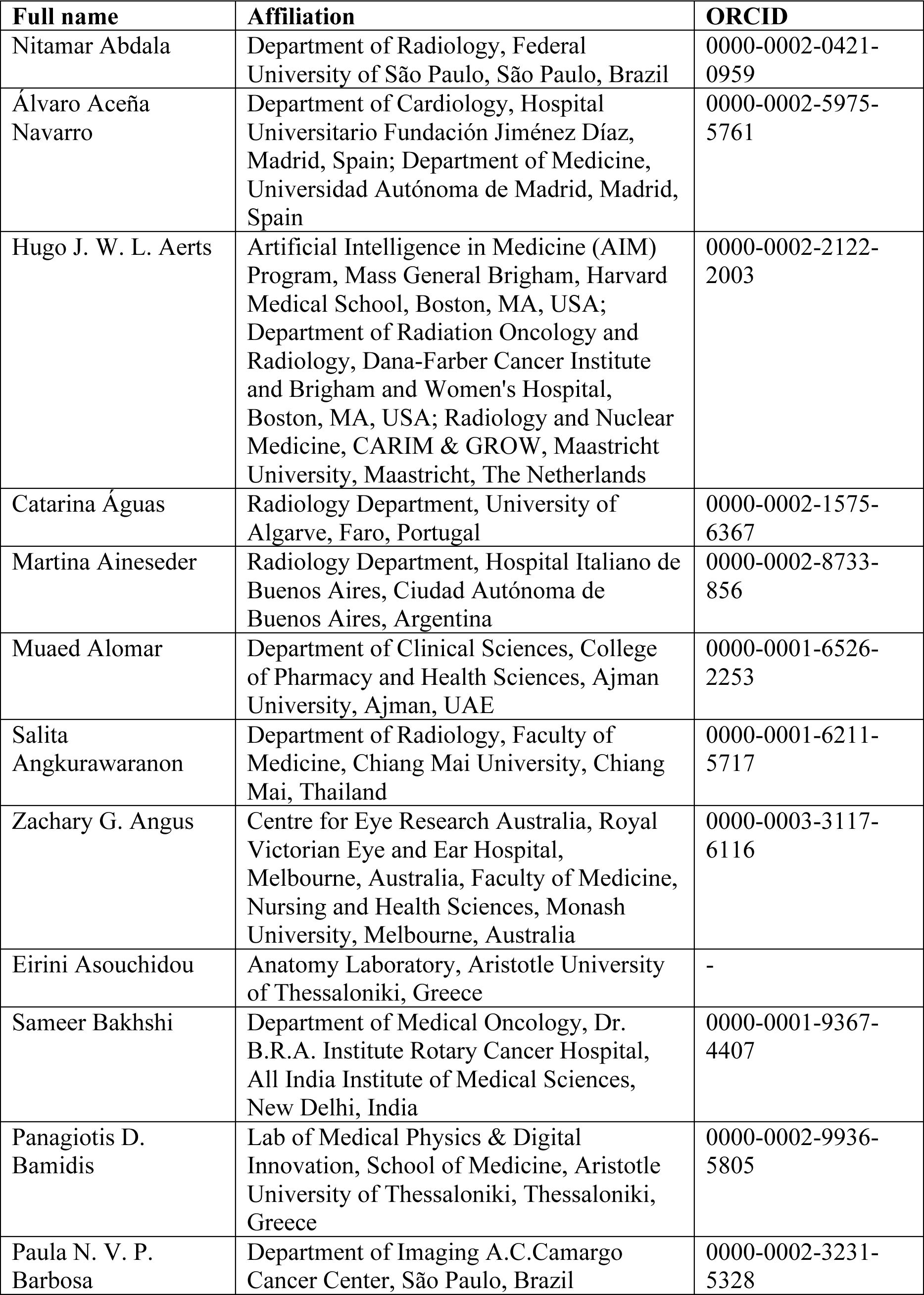
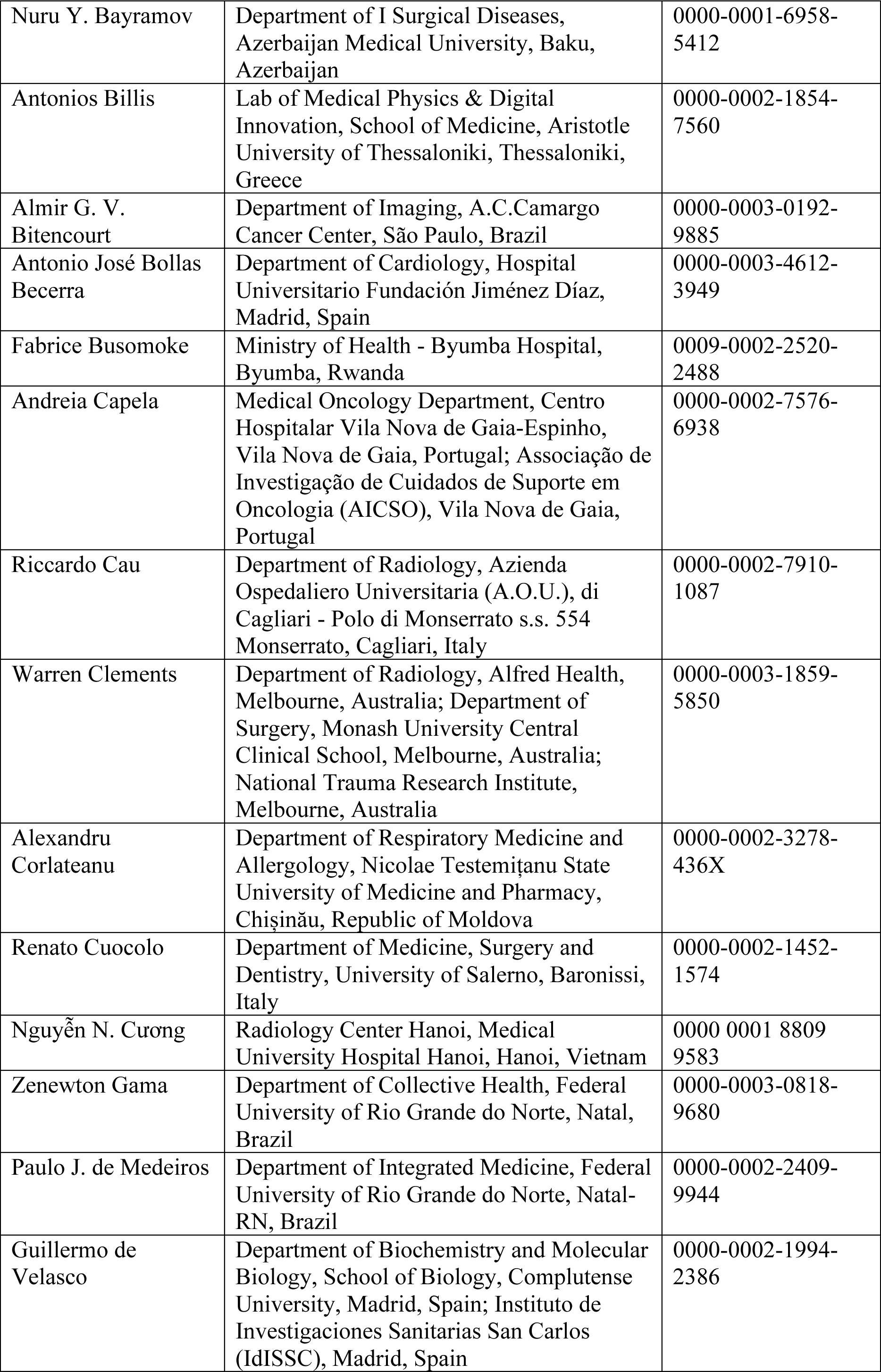
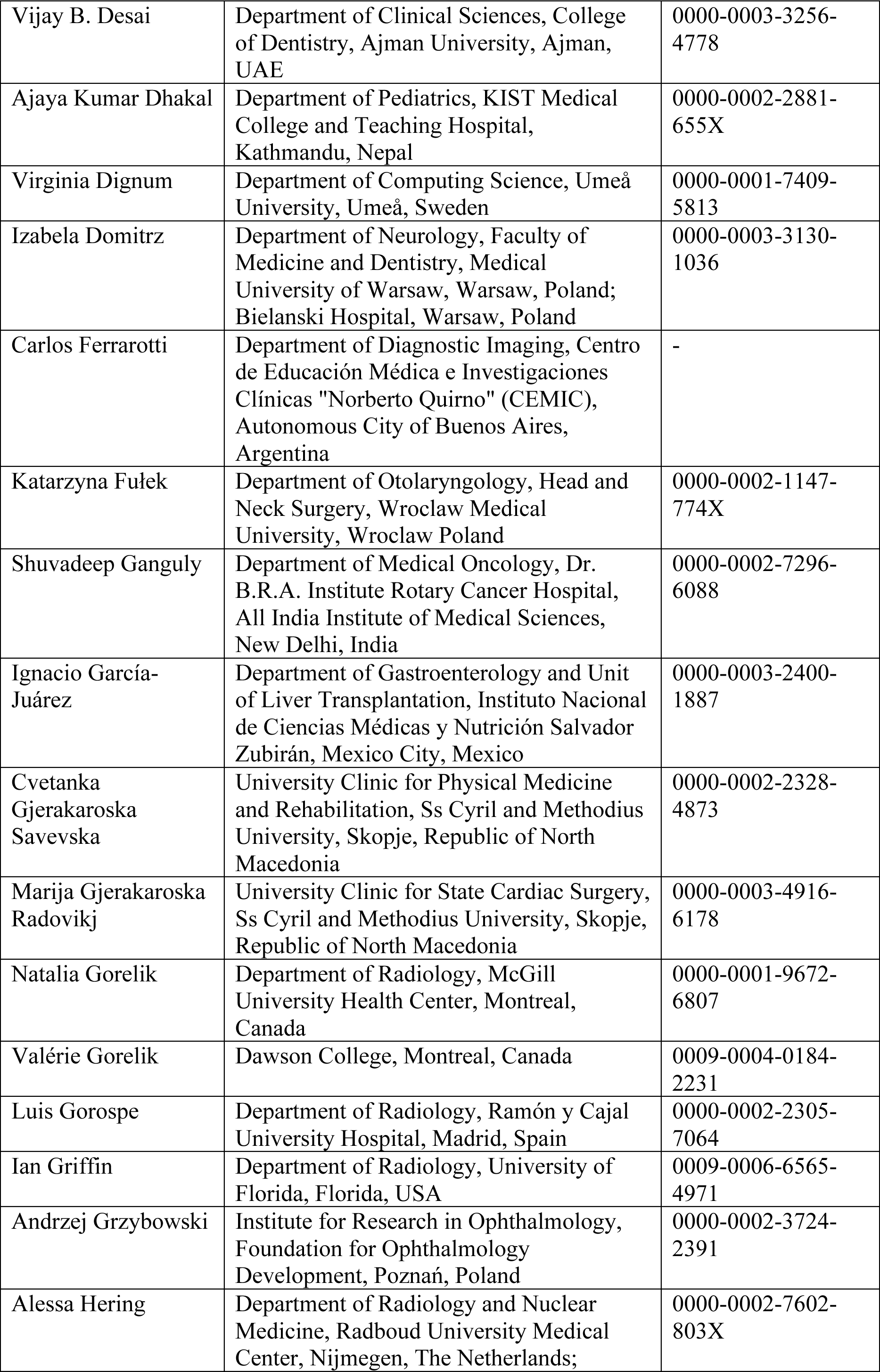
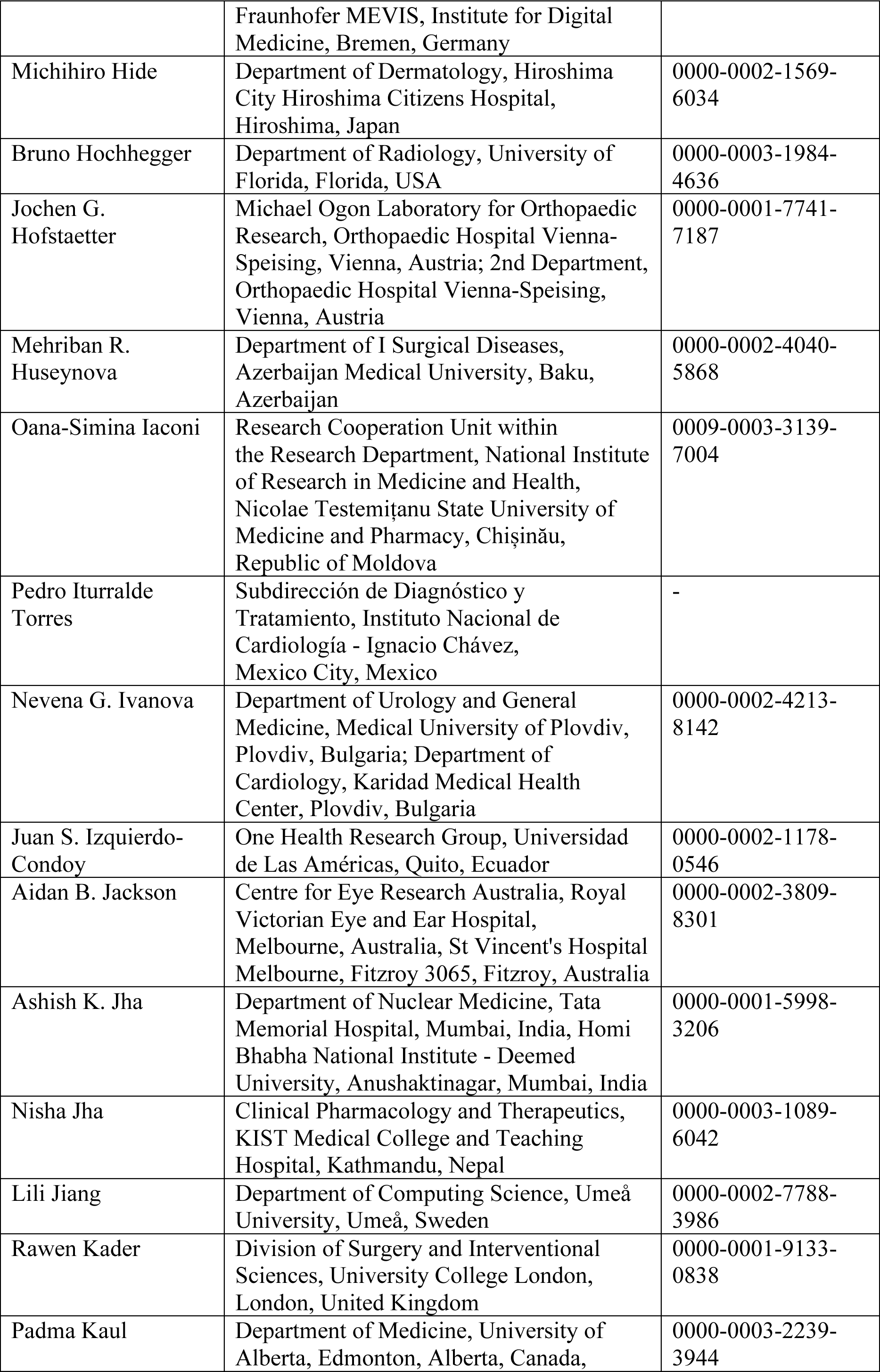
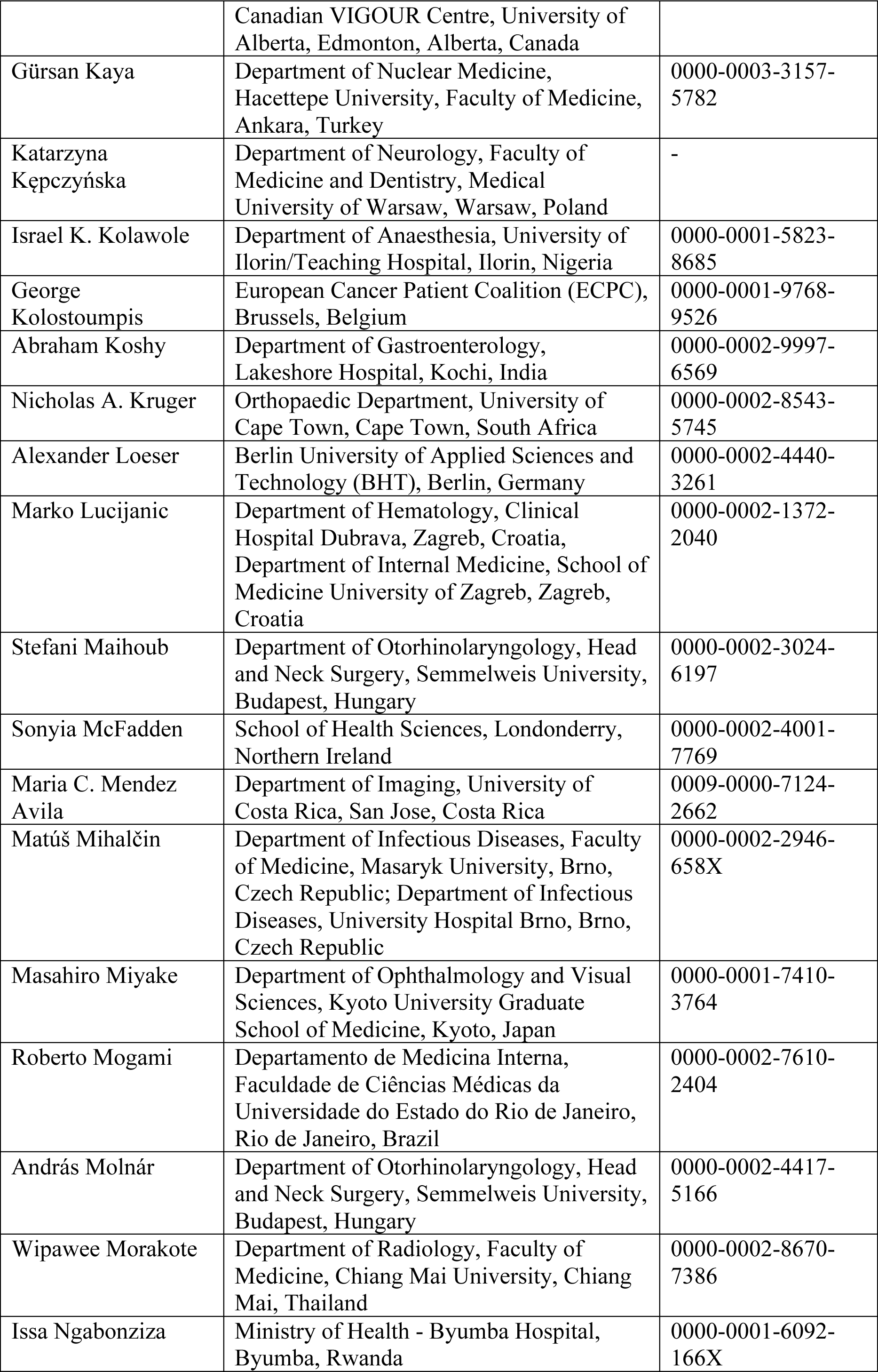
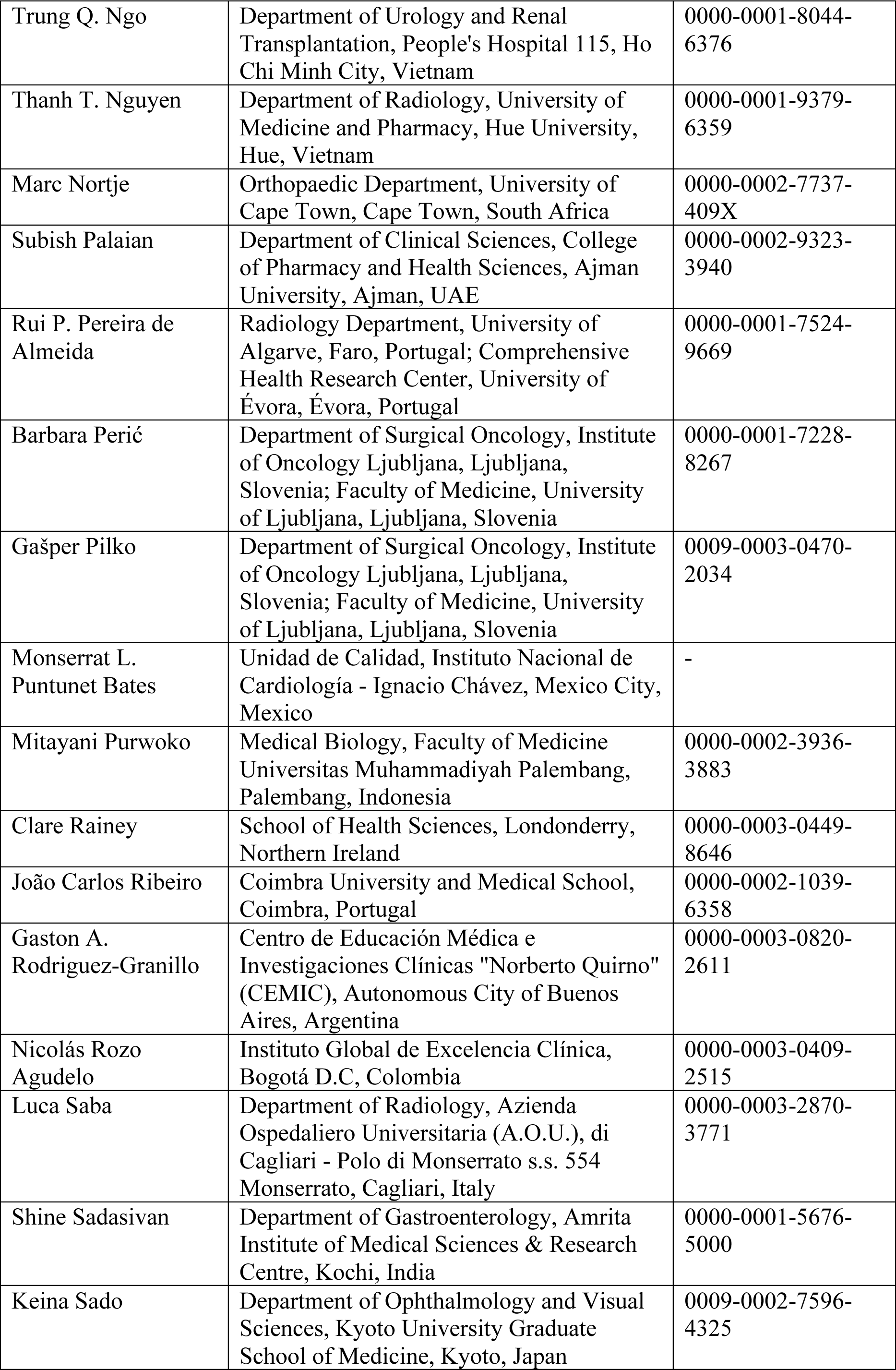
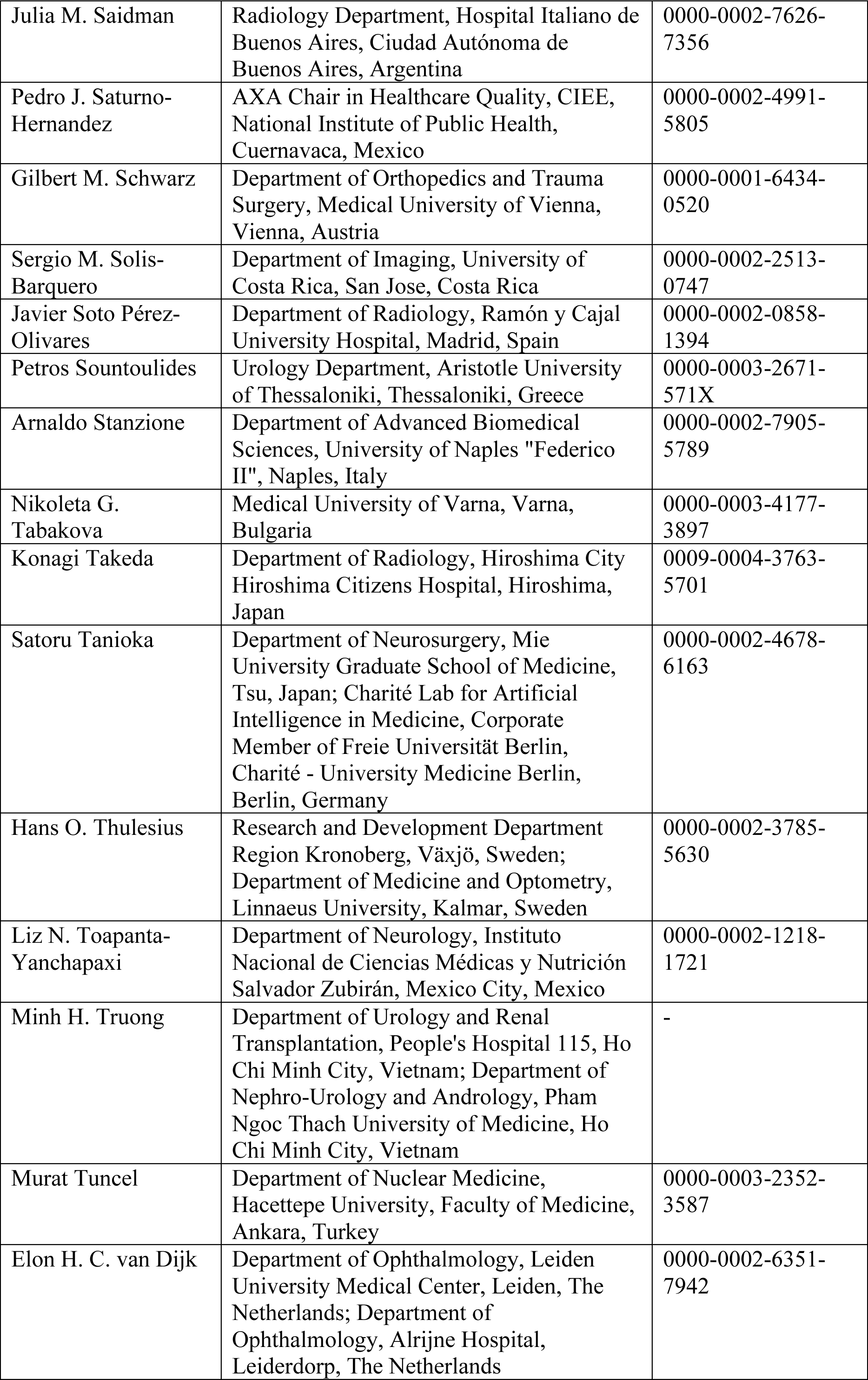
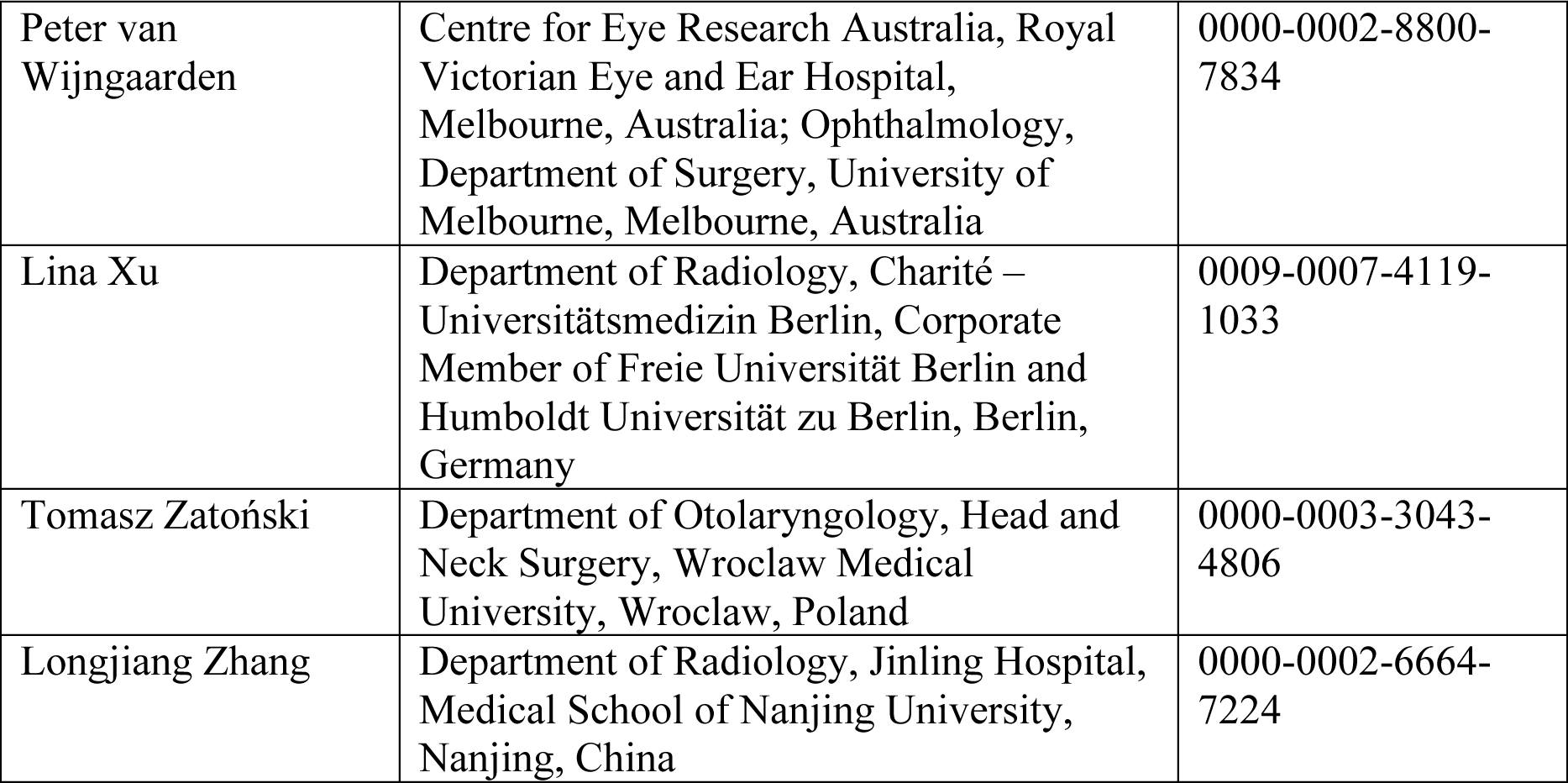

In addition, the authors want to thank Jaime Moujir-López, Javier Blázquez-Sánchez (Department of Radiology, Ramón y Cajal University Hospital, Madrid, Spain), Rubens Chojniak (Department of Imaging, A.C.Camargo Cancer Center, São Paulo, Brazil), and Dania Saad Rammal, Aya Mutasem Baradie, and Farrah Emad Elsubeihi (College of Pharmacy and Health Sciences, Ajman University, Ajman, United Arab Emirates) for supporting the data collection at their institutions. Keno K. Bressem is grateful for his participation in the Berlin Institute of Health (BIH) Charité Digital Clinician Scientist Program, funded by the Charité – Universitätsmedizin Berlin and the BIH.

## ICJME authorship contributions

**Conceptualization:** Felix Busch, Lena Hoffmann, Daniel Truhn, Marcus R. Makowski, Keno K. Bressem, Lisa C. Adams, Antonios Billis, Renato Cuocolo, Guillermo de Velasco, Luis Gorospe, Alessa Hering, Lili Jiang, Alexander Loeser, Javier Soto-Pérez- Olivares, Petros Sountoulides, Lina Xu; **Project administration:** Felix Busch, Lena Hoffmann, Keno K. Bressem, Lisa C. Adams; **Resources:** Felix Busch, Keno K. Bressem, Lisa C. Adams, COMFORT consortium; **Software:** Felix Busch, Lena Hoffmann, Keno K. Bressem, Lisa C. Adams; **Data curation:** Felix Busch, Lena Hoffmann, Keno K. Bressem, Lisa C. Adams; **Formal analysis:** Felix Busch, Lena Hoffmann, Keno K. Bressem, Lisa C. Adams, COMFORT consortium; **Funding acquisition:** Felix Busch, Marcus R. Makowski, Keno K. Bressem, Lisa C. Adams, Panagiotis D. Bamidis, Antonios Billis, Renato Cuocolo, Guillermo de Velasco, Virginia Dignum, Alessa Hering, Lili Jiang, George Kolostoumpis, Alexander Loeser, Petros Sountoulides; **Investigation:** Felix Busch, Lena Hoffmann, Keno K. Bressem, Lisa C. Adams; **Methodology:** Felix Busch, Lena Hoffmann, Daniel Truhn, Keno K. Bressem, Lisa C. Adams; **Supervision:** Felix Busch, Keno K. Bressem; **Validation:** Felix Busch, Lena Hoffmann, Daniel Truhn, Esteban Ortiz-Prado, Marcus R. Makowski, Keno K. Bressem, Lisa C. Adams, COMFORT consortium; **Visualization:** Felix Busch, Esteban Ortiz-Prado, Keno K. Bressem; **Writing – original draft preparation:** Felix Busch, Keno K. Bressem, Lisa C. Adams; **Writing – review & editing:** Felix Busch, Lena Hoffmann, Daniel Truhn, Esteban Ortiz-Prado, Marcus R. Makowski, Keno K. Bressem, Lisa C. Adams, COMFORT consortium.

All COMFORT consortia authors equally contributed to the data collection at their institutions, critically revised the final version of the manuscript for intellectual content, gave their final approval of the version to be published, and agreed to be accountable for all aspects of the work in ensuring that questions related to the accuracy or integrity of any part of the work are appropriately investigated and resolved.

## Data availability statement

Upon acceptance of the peer-reviewed article, the collected and analyzed dataset will be publicly available under CC-BY 4.0 license: Busch F, Hoffmann L, Truhn D, Ortiz-Prado E, Makowski MR, Bressem KK, et al. Dataset: Medical students’ perceptions towards artificial intelligence in education and practice: A multinational, multicenter cross-sectional study. Database: figshare [Internet]. doi:10.6084/m9.figshare.24422422 [Reserved]. For peer review, a private anonymized link is provided.

## Supporting information captions

S1 Table. Overview of survey participants divided by country, faculty, city, degree, number of enrolled students, and response rate.

S2 Table. Free-field comments by survey participants

S3 Table. Subgroup analysis by gender.

S4 Table. Subgroup analysis by age.

S5 Table. Subgroup analysis by study year.

S6 Table. Subgroup analysis by weekly use of technical devices.

S7 Table. Subgroup analysis by previous coding experience.

S8 Table. Subgroup analysis by AI knowledge.

S9 Table. Subgroup analysis by students who reported curricular events on AI in medicine of any duration versus those who indicated no curricular events

S10 Table. STROBE Statement—Checklist of items that should be included in reports of cross-sectional studies

