## Supporting information for "Medical students’ perceptions towards artificial intelligence in education and practice: A multinational, multicenter cross-sectional study"

^ Membership of the COMFORT Consortium is provided in the Acknowledgements of the corresponding manuscript.

### S1 Table. Overview of survey participants divided by country, faculty, city, degree, number of enrolled students, and response rate.

| Country/faculty | City | Degree (N) | | | Enrolled students (N) | Response rate (%) |
|---|---|---|---|---|---|---|
| | | (Human) Medicine | Dentistry | Veterinary Medicine | | |
| **Total** | | **4,313 (93.8%)** | **205 (4.5%)** | **78 (1.7%)** | **5,575,307** | **0.192 ± 0.445[a]** |
| **Argentina** | | **125 (3.0%)** | **0** | **0** | **362,089** | **0.140 ± 0.228[a]** |
| 1. Center for Clinical Medical Education and Research | Autonomous City of Buenos Aires | 18 | 0 | 0 | N/A | N/A |
| 2. Hospital Italiano de Buenos Aires | Autonomous City of Buenos Aires | 17 | 0 | 0 | N/A | N/A |
| 3. National University of Central Buenos Aires | Autonomous City of Buenos Aires | 45 | 0 | 0 | 11,142 | 0.404 |
| 4. National University of the Northeast | Corrientes | 1 | 0 | 0 | 53,308 | 0.002 |
| 5. University of Buenos Aires | Autonomous City of Buenos Aires | 44 | 0 | 0 | 297,639 | 0.015 |
| **Australia** | | **99 (2.4%)** | **1 (0.5%)** | **4 (5.1%)** | **152,614** | **0.064 ± 0.049[a]** |
| 6. Monash University | Melbourne | 47 | 0 | 0 | 58,460 | 0.080 |
| 7. University of Melbourne | Melbourne | 52 | 1 | 0 | 51,307 | 0.103 |
| 8. University of Queensland | Brisbane | 0 | 0 | 4 | 42,847 | 0.009 |
| **Austria** | | **155 (3.8%)** | **4 (2.0%)** | **0** | **8,871** | **1,386 ± 0.899[a]** |
| 9. Medical University Vienna | Vienna | 143 | 4 | 0 | 7,271 | 2.022 |
| 10. Paracelsus Medical University | Salzburg | 12 | 0 | 0 | 1,600 | 0.750 |
| **Azerbaijan** | | **100 (2.4%)** | **0** | **0** | **8,000** | **1,250 ± 0[a]** |
| 11. Azerbaijan Medical University | Baku | 100 | 0 | 0 | 8,000 | 1.250 |
| **Brazil** | | **235 (5.7%)** | **0** | **0** | **306,845** | **0.096 ± 0.144[a]** |
| 12. Federal University of Rio Grande do Norte | Natal | 149 | 0 | 0 | 42,906 | 0.347 |
| 13. Federal University of São Paulo | São Paulo | 41 | 0 | 0 | 83,182 | 0.049 |
| 14. Municipal University of São Caetano do Sul | São Caetano do Sul | 1 | 0 | 0 | 45,760 | 0.002 |
| 15. Rio de Janeiro State University | Rio de Janeiro | 39 | 0 | 0 | 52,534 | 0.074 |
| 16. University of Nove de Julho | São Paulo | 5 | 0 | 0 | 82,463 | 0.006 |
| **Bulgaria** | | **34 (0.8%)** | **0** | **0** | **22,083** | **0.185 ± 0.245[a]** |
| 17. Medical University of Sofia | Sofia | 1 | 0 | 0 | 10,849 | 0.009 |
| 18. Medical University of Varna | Varna | 29 | 0 | 0 | 6,234 | 0.465 |
| 19. University of Plovdiv | Plovdiv | 4 | 0 | 0 | 5,000 | 0.080 |
| **Canada** | | **107 (2.6%)** | **0** | **0** | **86,037** | **0.112 ± 0.096[a]** |
| 20. McGill University | Montreal | 58 | 0 | 0 | 30,821 | 0.188 |
| 21. University of Alberta | Edmonton | 48 | 0 | 0 | 33,730 | 0.142 |
| 22. University of Victoria | Victoria | 1 | 0 | 0 | 21,486 | 0.005 |
| **Chile** | | **71 (1.7%)** | **0** | **0** | **39,885** | **0.271 ± 0.257[a]** |
| 23. Catholic University of the Most Immaculate Conception | Concepción | 27 | 0 | 0 | 30,174 | 0.089 |
| 24. University of the Andes | Santiago de Chile | 44 | 0 | 0 | 9,711 | 0.435 |
| **China** | | **76 (1.8%)** | **0** | **0** | **125,945** | **0.048 ± 0.036[a]** |
| 25. Capital Medical University | Beijing | 5 | 0 | 0 | 15,347 | 0.033 |
| 26. Guangzhou Medical University | Guangzhou | 2 | 0 | 0 | 14,728 | 0.014 |
| 27. Lanzhou University | Lanzhou | 1 | 0 | 0 | N/A | N/A |
| 28. Nanjing University | Nanjing | 40 | 0 | 0 | 36,711 | 0.109 |
| 29. Shanghai Jiao Tong University School of Medicine | Shanghai | 22 | 0 | 0 | 38,472 | 0.057 |
| 30. Southern Medical University | Guangzhou | 4 | 0 | 0 | 6,575 | 0.061 |
| 31. Xuzhou Medical University | Xuzhou | 2 | 0 | 0 | 14,112 | 0.014 |
| **Colombia** | | **20 (0.5%)** | **0** | **0** | **N/A** | **N/A** |
| 32. Saint Martin University | Bogotá | 20 | 0 | 0 | N/A | N/A |
| **Costa Rica** | | **24 (0.6%)** | **11 (5.4%)** | **0** | **34,884** | **0.100 ± 0[a]** |
| 33. University of Costa Rica | San José | 24 | 11 | 0 | 34,884 | 0.100 |
| **Croatia** | | **10 (0.2%)** | **0** | **0** | **58,474** | **0.017 ± 0[a]** |
| 34. University of Zagreb | Zagreb | 10 | 0 | 0 | 58,474 | 0.017 |
| **Czech Republic** | | **31 (0.8%)** | **1 (0.5%)** | **0** | **32,789** | **0.098 ± 0[a]** |
| 35. Masaryk University | Brno | 31 | 1 | 0 | 32,789 | 0.098 |
| **Ecuador** | | **98 (2.4%)** | **0** | **6 (7.7%)** | **35,172** | **0.287 ± 0.346[a]** |
| 36. Central University of Ecuador | Quito | 21 | 0 | 0 | N/A | N/A |
| 37. Pontifical Catholic University of Ecuador | Quito | 9 | 0 | 0 | 21,237 | 0.042 |
| 38. University of Americans | Quito | 68 | 0 | 6 | 13,935 | 0.531 |
| **England** | | **38 (0.9%)** | **0** | **0** | **230,797** | **0.038 ± 0.057[a]** |
| 39. Barts and The London School of Medicine Dentistry | London | 6 | 0 | 0 | 3,410 | 0.176 |
| 40. Brighton and Sussex Medical School | Brighton | 1 | 0 | 0 | 977 | 0.102 |
| 41. Cambridge University | Cambridge | 1 | 0 | 0 | 20,565 | 0.005 |
| 42. Imperial College London | London | 3 | 0 | 0 | 20,275 | 0.015 |
| 43. University College London | London | 11 | 0 | 0 | 41,110 | 0.027 |
| 44. University of Bristol | Bristol | 1 | 0 | 0 | 27,335 | 0.004 |
| 45. University of Lincoln | Lincoln | 1 | 0 | 0 | 15,020 | 0.007 |
| 46. University of Manchester | Manchester | 2 | 0 | 0 | 40,725 | 0.005 |
| 47. University of Nottingham | Nottingham | 1 | 0 | 0 | 33,520 | 0.003 |
| 48. University of Sheffield | Sheffield | 11 | 0 | 0 | 27,860 | 0.039 |

| | | | | | | |
|---|---|---|---|---|---|---|
| **Ethiopia** | | **0** | **1 (0.5%)** | **0** | **N/A** | **N/A** |
| 49. Africa Medical College | Addis Ababa | 0 | 1 | 0 | N/A | N/A |
| **Germany** | | **675 (16.4%)** | **9 (4.4%)** | **61 (72.2%)** | **634,559** | **0.227 ± 0.370[a]** |
| 50. Charité – University Medicine Berlin | Berlin | 120 | 1 | 0 | 8,868 | 1.364 |
| 51. Free University of Berlin | Berlin | 0 | 0 | 11 | 25,427 | 0.043 |
| 52. Hanseatic University Rostock | Rostock | 126 | 6 | 1 | 14,000 | 0.950 |
| 53. Heidelberg University | Heidelberg | 8 | 0 | 0 | 19,315 | 0.041 |
| 54. Ludwig-Maximilians University of Munich | Munich | 22 | 0 | 0 | 34,622 | 0.064 |
| 55. Martin Luther University of Halle-Wittenberg | Halle-Wittenberg | 13 | 0 | 0 | 19,319 | 0.067 |
| 56. Philipps-University of Marburg | Marburg | 2 | 0 | 0 | 24,644 | 0.008 |
| 57. Ruhr University of Bochum | Bochum | 11 | 0 | 0 | 37,709 | 0.029 |
| 58. RWTH Aachen University | Aachen | 20 | 0 | 0 | 34,914 | 0.057 |
| 59. Saarland University | Saarland | 22 | 0 | 0 | 16,680 | 0.132 |
| 60. Technical University Munich | Munich | 6 | 0 | 0 | 35,506 | 0.017 |
| 61. Heinrich Heine University Düsseldorf | Düsseldorf | 13 | 0 | 0 | 28,121 | 0.046 |
| 62. University of Erlangen-Nuremberg | Erlangen-Nuremberg | 13 | 0 | 0 | 29,467 | 0.044 |
| 63. Goethe University of Frankfurt | Frankfurt | 2 | 0 | 0 | 28,572 | 0.007 |
| 64. University of Freiburg | Freiburg | 9 | 0 | 0 | 19,417 | 0.046 |
| 65. Justus Liebig University of Giessen | Giessen | 0 | 0 | 14 | 27,184 | 0.052 |
| 66. University of Greifswald | Greifswald | 1 | 0 | 0 | 10,366 | 0.010 |
| 67. University of Köln | Köln | 1 | 0 | 0 | 31,299 | 0.030 |
| 68. University of Leipzig | Leipzig | 37 | 0 | 8 | 28,275 | 0.159 |
| 69. University of Lübeck | Lübeck | 25 | 0 | 0 | 3,921 | 0.638 |
| 70. Johannes Gutenberg University of Mainz | Mainz | 9 | 2 | 0 | 30,755 | 0.036 |
| 71. University of Münster | Münster | 75 | 0 | 0 | 27,529 | 0.272 |
| 72. University of Oldenburg | Oldenburg | 6 | 0 | 0 | 16,244 | 0.037 |
| 73. University of Regensburg | Regensburg | 77 | 0 | 0 | 20,321 | 0.379 |
| 74. University of Tübingen | Tübingen | 1 | 0 | 0 | 21,632 | 0.005 |
| 75. University of Ulm | Ulm | 15 | 0 | 1 | 10,301 | 0.155 |
| 76. University of Medicine Hannover | Hannover | 0 | 0 | 26 | 3,362 | 0.773 |
| 77. University of Witten/Herdecke | Witten/Herdecke | 35 | 0 | 0 | 3,152 | 1.110 |
| 78. University of Würzburg | Würzburg | 6 | 0 | 0 | 23,637 | 0.025 |
| **Greece** | | **36 (0.9%)** | **0** | **0** | **49,654** | **0.073 ± 0[a]** |
| 79. School of Medicine, Aristotle University of Thessaloniki | Thessaloniki | 36 | 0 | 0 | 49,654 | 0.073 |
| **Hungary** | | **44 (1.1%)** | **5 (2.4%)** | **0** | **9,966** | **0.492 ± 0[a]** |
| 80. Semmelweis University | Budapest | 44 | 5 | 0 | 9,966 | 0.492 |
| **India** | | **137 (3.3%)** | **0** | **0** | **25,294** | **1.786 ± 2.366[a]** |
| 81. All India Institute of Medical Sciences | New Delhi | 111 | 0 | 0 | 3,209 | 3.459 |
| 82. Amrita Vishwa Vidyapeetham | Kochi | 25 | 0 | 0 | 22,085 | 0.113 |
| 83. Tata Memorial Hospital | Mumbai | 1 | 0 | 0 | N/A | N/A |
| **Indonesia** | | **124 (3.0%)** | **0** | **0** | **12,000** | **1.033 ± 0[a]** |
| 84. University Muhammadiyah Palembang | Palembang | 124 | 0 | 0 | 12,000 | 1.033 |
| **Ireland** | | **2 (<0.0%)** | **0** | **0** | **38,045** | **0.006 ± 0.002[a]** |
| 85. University of Limerick | Limerick | 1 | 0 | 0 | 14,640 | 0.007 |
| 86. University College Dublin | Dublin | 1 | 0 | 0 | 23,405 | 0.004 |
| **Italy** | | **141 (3.4%)** | **2 (1.0%)** | **1 (1.3%)** | **230,102** | **0.108 ± 0.146[a]** |
| 87. Sapienza University of Rome | Rome | 2 | 0 | 0 | 60,522 | 0.003 |
| 88. University of Cagliari | Cagliari | 76 | 2 | 0 | 26,028 | 0.300 |
| 89. University of Milano | Milano | 1 | 0 | 0 | 61,283 | 0.002 |
| 90. University of Naples Federico II | Naples | 2 | 0 | 0 | 56,127 | 0.004 |
| 91. University of Salerno | Salerno | 60 | 0 | 1 | 26,142 | 0.233 |
| **Japan** | | **88 (2.1%)** | **13 (6.3%)** | **0** | **201,571** | **0.051 ± 0.086[a]** |
| 92. Chiba University | Chiba | 1 | 0 | 0 | 13,832 | 0.007 |
| 93. Ehime University | Matsuyama | 3 | 0 | 0 | 9,101 | 0.033 |
| 94. Hiroshima University | Hiroshima | 24 | 13 | 0 | 14,590 | 0.254 |
| 95. Hyogo Medical University | Nishinomiya | 1 | 0 | 0 | 2,603 | 0.038 |
| 96. Juntendo University | Tokyo | 1 | 0 | 0 | 6,579 | 0.015 |
| 97. Kagawa University | Takamatsu | 1 | 0 | 0 | 6,309 | 0.016 |
| 98. Kagoshima University | Kagoshima | 1 | 0 | 0 | 10,577 | 0.009 |
| 99. Kanazawa University | Kanazawa | 1 | 0 | 0 | 10,271 | 0.010 |
| 100. Kitasato University | Tokyo | 1 | 0 | 0 | 8,628 | 0.012 |
| 101. Kumamoto University | Kumamoto | 1 | 0 | 0 | 9,384 | 0.011 |
| 102. Kyoto University | Kyoto | 7 | 0 | 0 | 21,817 | 0.032 |
| 103. Kyushu University | Fukuoka | 1 | 0 | 0 | 18,220 | 0.005 |
| 104. Mie University | Tsu | 14 | 0 | 0 | 6,983 | 0.200 |
| 105. Nagoya University | Nagoya | 1 | 0 | 0 | 15,224 | 0.007 |
| 106. Oita University | Oita | 1 | 0 | 0 | 5,381 | 0.019 |
| 107. Okayama University | Okayama | 3 | 0 | 0 | 13,027 | 0.023 |
| 108. Saga University | Saga | 2 | 0 | 0 | 6,518 | 0.031 |
| 109. Shimane University | Matsue | 1 | 0 | 0 | 6,005 | 0.017 |
| 110. Tokyo Medical and Dental University | Tokyo | 1 | 0 | 0 | 2,990 | 0.033 |
| 111. Tottori University | Tottori | 1 | 0 | 0 | 5,998 | 0.017 |
| 112. University of the Ryukyus | Nishihara | 21 | 0 | 0 | 7,534 | 0.279 |
| **Kosovo** | | **1 (<0.0%)** | **0** | **0** | **28,532** | **0.004 ± 0[a]** |
| 113. University for Business and Technology | Prishtina | 1 | 0 | 0 | 28,532 | 0.004 |
| **Mexico** | | **317 (7.7%)** | **0** | **4 (5.1%)** | **1,192,919** | **0.055 ± 0.167[a]** |
| 114. Autonomous University of Aguascalientes | Aguascalientes | 1 | 0 | 0 | 14,606 | 0.007 |
| 115. Autonomous University of Baja California | Mexicali | 4 | 0 | 0 | 67,944 | 0.006 |
| 116. Autonomous University of Chihuahua | Chihuahua | 6 | 0 | 0 | 25,302 | 0.024 |
| 117. Autonomous University of Durango | Durango City | 14 | 0 | 0 | 16,036 | 0.087 |
| 118. Autonomous University of Guadalajara | Guadalajara | 1 | 0 | 0 | 140,348 | 0.001 |

| | | | | | | |
|---|---|---|---|---|---|---|
| 119. Autonomous University of Guerrero | Chilpancingo de los Bravo | 1 | 0 | 0 | 35,448 | 0.003 |
| 120. Autonomous University of Nayarit | Tepic | 4 | 0 | 0 | 26,501 | 0.015 |
| 121. Autonomous University of Sinaloa | Culiacán | 2 | 0 | 0 | 74,001 | 0.003 |
| 122. Autonomous University of Yucatán | Mérida | 1 | 0 | 0 | 19,128 | 0.005 |
| 123. Autonomous University of Zacatecas | Zacatecas | 1 | 0 | 0 | 28,311 | 0.004 |
| 124. Cuauhtémoc University | Puebla | 3 | 0 | 0 | 12,437 | 0.024 |
| 125. Universidad La Salle | Mexico City | 2 | 0 | 0 | 9,094 | 0.022 |
| 126. Meritorious Autonomous University of Puebla | Puebla | 15 | 0 | 0 | 85,321 | 0.018 |
| 127. Michoacan University of Saint Nicholas of Hidalgo | Morelia | 23 | 0 | 0 | 47,425 | 0.048 |
| 128. National Autonomous University of Mexico | Mexico City | 87 | 0 | 0 | 175,561 | 0.050 |
| 129. National Polytechnic Institute | Mexico City | 3 | 0 | 0 | 166,738 | 0.002 |
| 130. Universidad Pablo Guardado Chávez | Tuxtla Gutiérrez | 1 | 0 | 0 | 2,974 | 0.034 |
| 131. University Del Valle de Mexico | Mexico City | 17 | 0 | 3 | 33,442 | 0.060 |
| 132. University of Guanajuato | Guanajuato | 1 | 0 | 0 | 30,317 | 0.003 |
| 133. University of Monterrey | Monterrey | 96 | 0 | 1 | 11,670 | 0.831 |
| 134. University of Quintana Roo | Chetumal | 1 | 0 | 0 | 4,320 | 0.023 |
| 135. University of Sonora | Hermosillo | 3 | 0 | 0 | 40,995 | 0.007 |
| 136. University of Vasco de Quiroga | Morelia | 2 | 0 | 0 | 45,000 | 0.004 |
| 137. University of Veracruz | Xalapa | 28 | 0 | 0 | 80,000 | 0.035 |
| **Nepal** | | **47 (1.1%)** | **8 (3.9%)** | **0** | **482,541** | **0.011 ± 0[a]** |
| 138. Tribhuvan University | Kathmandu | 47 | 8 | 0 | 482,541 | 0.011 |
| **Nigeria** | | **51 (1.2%)** | **0** | **0** | **85,710** | **0.042 ± 0.038[a]** |
| 139. Bowen University | Iwo | 1 | 0 | 0 | 4,500 | 0.022 |
| 140. Ladoke Akintola University of Technology | Ogbomoso | 5 | 0 | 0 | 28,289 | 0.018 |
| 141. University of Ilorin | Ilorin | 45 | 0 | 0 | 52,921 | 0.085 |
| **Northern Ireland** | | **1 (<0.0%)** | **1 (0.5%)** | **0** | **19,825** | **0.010 ± 0[a]** |
| 142. Ulster University | Belfast | 1 | 1 | 0 | 19,825 | 0.010 |
| **Poland** | | **227 (5.5%)** | **3 (1.5%)** | **1 (1.3%)** | **110,274** | **0.236 ± 0.385[a]** |
| 143. Jagiellonian University Medical College | Kraków | 1 | 0 | 0 | 34,309 | 0.003 |
| 144. Medical University of Gdansk | Gdańsk | 2 | 0 | 0 | 6,340 | 0.032 |
| 145. Medical University of Lodz | Lodz | 1 | 0 | 0 | 10,237 | 0.010 |
| 146. Medical University of Lublin | Lublin | 1 | 0 | 0 | 6,884 | 0.015 |
| 147. Medical University of Silesia | Katowice | 1 | 0 | 0 | 10,237 | 0.010 |
| 148. Medical University of Warsaw | Warsaw | 32 | 3 | 0 | 10,068 | 0.348 |
| 149. Poznań University of Medical Sciences | Poznań | 0 | 0 | 1 | 7,296 | 0.014 |
| 150. University of Warmia and Mazury | Olsztyn | 101 | 0 | 0 | 16,889 | 0.598 |
| 151. Wroclaw Medical University | Wroclaw | 88 | 0 | 0 | 8,014 | 1.098 |
| **Portugal** | | **332 (8.1%)** | **0** | **0** | **129,441** | **0.245 ± 0.123[a]** |
| 152. University of Algarve | Faro | 18 | 0 | 0 | 8,878 | 0.203 |
| 153. University of Beira Interior | Covilhã | 38 | 0 | 0 | 8,248 | 0.461 |
| 154. University of Coimbra | Coimbra | 36 | 0 | 0 | 21,767 | 0.165 |
| 155. University of Lisbon | Lisbon | 165 | 0 | 0 | 49,847 | 0.331 |
| 156. University of Porto | Porto | 70 | 0 | 0 | 36,386 | 0.192 |
| 157. University of the Azores | Ponta Delgada | 5 | 0 | 0 | 4,315 | 0.116 |
| **Republic of Moldova** | | **67 (1.6%)** | **0** | **0** | **7,000** | **0.957 ± 0[a]** |
| 158. Nicolae Testemitanu State University of Medicine and Pharmacy | Chişinău | 67 | 0 | 0 | 7,000 | 0.957 |
| **Republic of North Macedonia** | | **120 (2.9%)** | **3 (1.5%)** | **0** | **32,317** | **0.370 ± 0.026[a]** |
| 159. University of Ss. Cyril and Methodius | Skopje | 96 | 2 | 0 | 25,220 | 0.389 |
| 160. University of Tetova | Tetovo | 24 | 1 | 0 | 7,097 | 0.352 |
| **Romania** | | **1 (<0.0%)** | **0** | **0** | **9,258** | **0.011 ± 0[a]** |
| 161. University of Medicine, Pharmacy, Science and Technology of Târgu Mureş | Târgu Mureş | 1 | 0 | 0 | 9,258 | 0.011 |
| **Russia** | | **0** | **1 (0.5%)** | **0** | **N/A** | **N/A** |
| 162. N/A | N/A | 0 | 1 | 0 | N/A | N/A |
| **Rwanda** | | **4 (0.1%)** | **0** | **0** | **52,000** | **0.008 ± 0[a]** |
| 163. Mount Kenya University | Thika | 4 | 0 | 0 | 52,000 | 0.008 |
| **Slovenia** | | **87 (2.1%)** | **19 (9.3%)** | **0** | **53,526** | **0.141 ± 0.156[a]** |
| 164. University of Ljubljana | Ljubljana | 84 | 18 | 0 | 40,607 | 0.251 |
| 165. University of Maribor | Maribor | 3 | 1 | 0 | 12,919 | 0.031 |
| **South Africa** | | **63 (1.5%)** | **0** | **0** | **47,101** | **0.143 ± 0.191[a]** |
| 166. University of Cape Town | Cape Town | 61 | 0 | 0 | 21,961 | 0.278 |
| 167. University of Stellenbosch | Stellenbosch | 2 | 0 | 0 | 25,140 | 0.008 |
| **Spain** | | **74 (1.8%)** | **0** | **0** | **79,403** | **0.073 ± 0.090[a]** |
| 168. Autonomous University of Madrid | Madrid | 37 | 0 | 0 | 30,277 | 0.122 |
| 169. CEU San Pablo University | Madrid | 1 | 0 | 0 | 8,138 | 0.012 |
| 170. University Francisco de Vitoria | Madrid | 1 | 0 | 0 | 5,500 | 0.018 |
| 171. University of Alcalá | Madrid | 34 | 0 | 0 | 16,260 | 0.209 |
| 172. University of La Laguna | San Cristóbal de La Laguna | 1 | 0 | 0 | 19,228 | 0.005 |
| **Sweden** | | **30 (0.7%)** | **1 (0.5%)** | **0** | **34,006** | **0.092 ± 0.018[a]** |
| 173. Linköping University | Linköping | 16 | 0 | 0 | 15,233 | 0.105 |
| 174. Umeå University | Umeå | 14 | 1 | 0 | 18,773 | 0.080 |
| **Switzerland** | | **119 (2.9%)** | **1 (0.5%)** | **0** | **53,468** | **0.248 ± 0.149[a]** |
| 175. Università della Svizzera Italiana | Lugano | 13 | 0 | 0 | 3,714 | 0.350 |
| 176. University of Basel | Basel | 28 | 0 | 0 | 8,536 | 0.328 |
| 177. University of Bern | Bern | 47 | 1 | 0 | 12,399 | 0.387 |
| 178. University of Lucerne | Lucerne | 2 | 0 | 0 | 3,211 | 0.062 |
| 179. University of Zurich | Zurich | 29 | 0 | 0 | 25,608 | 0.113 |

| | | | | | | |
|---|---|---|---|---|---|---|
| **Tanzania** | | **6 (0.1%)** | **0** | **0** | **1,100** | **0.545 ± 0[a]** |
| 180. Kilimanjaro Christian Medical University College | Moshi | 6 | 0 | 0 | 1,100 | 0.545 |
| **Thailand** | | **49 (1.2%)** | **0** | **0** | **33,649** | **0.146 ± 0[a]** |
| 181. Chiang Mai University | Chiang Mai | 49 | 0 | 0 | 33,649 | 0.146 |
| **The Netherlands** | | **13 (0.3%)** | **0** | **0** | **35,072** | **0.037 ± 0[a]** |
| 182. Leiden University | Leiden | 13 | 0 | 0 | 35,072 | 0.037 |
| **Turkey** | | **36 (0.9%)** | **1 (0.5%)** | **0** | **37,249** | **0.099 ± 0[a]** |
| 183. Hacettepe University | Ankara | 36 | 1 | 0 | 37,249 | 0.099 |
| **United Arab Emirates** | | **45 (1.1%)** | **117 (57.1%)** | **1 (1.3%)** | **4,823** | **3.380 ± 0[a]** |
| 184. Ajman University | Ajman | 45 | 117 | 1 | 4,823 | 3.380 |
| **United States of America** | | **52 (1.3%)** | **0** | **0** | **106,275** | **0.071 ± 0.077[a]** |
| 185. Duke University | Durham | 24 | 0 | 0 | 15,527 | 0.155 |
| 186. University of California | Oakland | 1 | 0 | 0 | 42,634 | 0.002 |
| 187. University of Florida | Gainesville | 27 | 0 | 0 | 48,114 | 0.056 |
| **Vietnam** | | **101 (2.5%)** | **3 (1.5%)** | **0** | **234,142** | **0.042 ± 0.060[a]** |
| 188. Ho Chi Minh City Medicine and Pharmacy University | Ho Chi Minh City | 2 | 0 | 0 | 83,599 | 0.002 |
| 189. Hue University of Medicine and Pharmacy | Hue | 61 | 3 | 0 | 48,943 | 0.131 |
| 190. Pham Ngoc Thach University of Medicine | Ho Chi Minh City | 13 | 0 | 0 | N/A | N/A |
| 191. Tra Vinh University | Tra Vinh | 1 | 0 | 0 | 11,600 | 0.009 |
| 192. Vietnam Military Medical University | Hanoi | 22 | 0 | 0 | 90,000 | 0.024 |
| **N/A** | **N/A** | **2 (<0.0%)** | **0** | **0** | **N/A** | **N/A** |

The total number of students enrolled in each faculty was determined using the official webpage, if available, or the Times Higher Education World University Rankings 2024.

[a] Reported as mean ± standard deviation.

N/A, not available.

### S2 Table. Free-field comments by survey participants.

| Comment | |
|---|---|
| 1. | Setting early ethical regulation measures at congresses and meetings of all medical areas focused on AI. |
| 2. | Even if AI is available, right now, the implanted technology in the hospitals is way too bad to benefit from AI at all because it only causes more work. |
| 3. | No need for AI education. |
| 4. | It would be nice if medical universities would arrive in the 21st century. |
| 5. | I'm not representative as I already have a degree in Bioinformatics. |
| 6. | Regarding the question, "Suppose an artificial intelligence (AI) makes a diagnosis. What would you prefer?" -> It depends on how often people have the condition being tested for (prior probability, thinking of Bayes rule here - that is something we should learn about more, actually) |
| 7. | In my opinion, it is not understandable why AI is still no topic in human medicine university teaching. It has the potential to be a powerful tool in healthcare but needs to be handled with great care. For this reason, tight cooperation in development between software developers, universities, and healthcare professionals is urgently needed. |
| 8. | Please, no new programming courses in the following years. |
| 9. | I think AI can't replace medical workers, and we shouldn't make it seem like AI is better than humans... |
| 10. | Artificial intelligence has been of great help not just to students and doctors but also to patients. Its use in the future would be a great help for the betterment and holistic development of a healthy society. |
| 11. | I would love to work with AI. |
| 12. | In Japan and Singapore, they are working on making AI machines for surgery. |
| 13. | AI is a rather helpful tool, but in my opinion, it couldn't replace physicians. Physicians should not base their future diagnosis only on AI or similar tools like ChatGPT; it is in the best interest to provide future education on how they could help physicians and their patients. |
| 14. | Like every other technology built by humans, everything has positive and negative aspects; AI technology is still rather new and unexplored to its full potential, which is why it may be difficult to interpret its exact importance in the medical field accurately for the future. Nevertheless, I believe it is the next important step in technological development in order to improve in the future. |
| 15. | AI is the future of medicine. |
| 16. | I had two elective courses on AI, but from the University of Rijeka (before I transferred to Ljubljana), so my answers are more tied to that university. |
| 17. | I believe AI to be very useful, but I don't expect it to be used often in primary or secondary care due to fear and lack of knowledge about it, especially from doctors, not as much from patients. |
| 18. | The most important skill for physicians using AI could be the ability to assess the accuracy/the source of the AI knowledge. |
| 19. | AI would never be smarter than people because it is something artificial. |
| 20. | AI should be supported but not be dependent upon it. |
| 21. | I'm excited to think that AI will be a useful tool, but afraid that it may replace us. |
| 22. | AI must be developed in controlled settings, or humankind will lose control of this power. |
| 23. | I think the rather rigid structure at Uni hospitals will delay the implementation and use. |
| 24. | I believe that AI could be beneficial for diagnosis (e.g., evaluating an x-ray) but should not (at some point) replace a physician because I believe that there is more to a good healing process than just the medical treatment in itself (e.g., patient-physician interaction, real empathy, human charisma). |
| 25. | AI may only be used as a secondary tool. |
| 26. | I strongly believe that the implementation of AI in medicine will greatly negatively affect patient care as it will lower the workload of doctors, making them obsolete and completely removing the human element in making hard and quick decisions that come with saving a human life. |
| 27. | AI and VR could be amazing teaching methods. |
| 28. | Artificial intelligence is a great deal for the future of many fields. Medicine should not be left behind in this advancement. |
| 29. | I recommend AI. |
| 30. | Artificial intelligence has no emotions or empathy to treat humans, but it can be used for practicals in skill labs for studying and learning purposes. |
| 31. | A lot depends on how the physician's decision-making depends on the AI-submitted answers. Therefore, the physician should be well-trained to comprehend the dimensions and, more importantly, the limits of AI. |
| 32. | Medical students often lack basic concepts of IT/CS, so I fear that we'll be using it in the future without a deeper understanding of how the results came to happen. Star Trek would be an obvious idea, but I fear that insurance companies will use AI to determine if a treatment is worth it for a patient. |
| 33. | I think human contact is important. AIs might be efficient for organizations, but they should not make decisions by themselves without a human checking on them. |
| 34. | It is important to keep up with the times and to work efficiently but also competently in the interest of the patient. AI can be an advantage in this! |
| 35. | I don't think that AI can replace a real doctor, but it can help find diagnoses and build a network between countries and continents where difficult and rare illnesses can be shared. |
| 36. | I think the biggest problems with AI are if doctors just listen to it instead of making their own opinions and if the AI makes mistakes or gives false information. |
| 37. | We tried out ChatGPT in a pharmacy course and spotted some very bad medical decisions made by AI. That's why I am very careful with using AI for medical decisions and questions. |
| 38. | I think that it is important to implement an AI that is specifically designed for medicine. That communicates on a level that only those who study medicine can understand it because if it is explained too simply, it can lead to false assumptions. |
| 39. | I am afraid that people will use AI themselves to diagnose at home and will miss, therefore, adequate medical care. |
| 40. | I think that the cooperation of physicians and AI will be quite advantageous. |
| 41. | AI will be able to help with a lot of small tasks, giving the doctors more time to attend to more patients or get more rest so they aren't overworked. |
| 42. | We need to implement AI into medical school programs, not only the medical profession! It must be a student's assistant during medical school. |
| 43. | AI is good for medicine but with caution. |
| 44. | AI has to be a tool for humans and can't substitute it. Imagine a world where an AI decides all treatments. I would feel terrible as a patient. |

| | |
|---|---|
| 45. | AI may assist physicians to have a good direction in finding diagnosis while saving valuable time; therefore, more patients will get the proper treatment on time. |
| 46. | I think there is a high chance AI will be used as an aid rather than have a role in which it will be giving out diagnoses. I believe there will be a long way to go before people start to trust AI, and it will need to be used as an auxiliary first, introducing it gradually so that it can become more well-regulated before it has such an impact on patients' healthcare (i.e., discussing what the treatment will be for a newly diagnosed disease). |
| 47. | I would like to learn about artificial intelligence in general fields, especially its benefits for medicine. |
| 48. | It would be great if today's health world could be introduced to AI. |
| 49. | AI is the next necessary step in medical advancement, in my opinion. |
| 50. | AI is good, with some limits. |
| 51. | Technology is taking over! |
| 52. | I am afraid that people will use AI as doctors without consulting with a real one. |
| 53. | Great initiative! |
| 54. | The physicians will be needed yet. |
| 55. | I am very happy to participate in this revolutionary field of study! |
| 56. | Please incorporate this into our curriculum. There's a lot more to medicine than just biology, and we need the perspective of new developments shared separately and with the importance that they are due. |
| 57. | A great survey. I hope for the success and impact of your research. |
| 58. | AI should be taught from the school days. |
| 59. | Above all, I would like to know how AI can be a tool in radiology. |
| 60. | I would like to share a confusion that frequently pops into my mind. Would our 10-year training have that much value, or would it be easy to replace a doctor with some two-month trained personnel as the accuracy of the programs increases? I feel we, as students, are confused about our future role when AI is introduced. |
| 61. | I think AI would be better to use for situations where you don't interact with the patient, as most AI would predictably be much less empathetic. AI would be incredibly useful for Clinical Geneticists when doing lab tests with DNA. |
| 62. | ChatGPT is really awesome and can help me with my tasks in med school. |
| 63. | I view AI as a tool like AMBOSS or Google, not as a substitute for my medical knowledge. |
| 64. | I fear that in Germany, the implementation of medical procedures will still take a long time due to so many regulations. |
| 65. | In my opinion, AI can be supportive in diagnosis and treatment, but the last decision should always be made by the physician. Human beings and biology do not always use the same pathways. |
| 66. | I think a combination of AI and learned knowledge to prove AI diagnosis is necessary. |
| 67. | There is a need for more professorships on AI in medicine. |
| 68. | I work in clinical research in the area of AI. |
| 69. | Personally, I don't think AI could ever replace experience and that kind of gut feeling with a situation or patient. So, therefore, I can't imagine a symbiosis in our future everyday work. It's rather helpful with other tasks, I guess, especially during lectures or exam situations. |
| 70. | More teaching on how to use AI and how to implement it in daily usage. |
| 71. | Autopilot in aviation was not so difficult that we didn't need machine learning. The structure of using machine learning in medicine is similar, but semi-automating medicine requires extreme design consideration. |
| 72. | I think that using AI in medicine is a good idea itself, but what I'm worried about is that people always tend to misuse good things. |
| 73. | Medicine is great, but our studies lack practice. |
| 74. | Never again, appointments in the paper calendar. I want the 21st century. AI knows better anyway when I have time in 6 years. |
| 75. | AI should work with modes for you to choose from: high sensitivity mode, high specificity mode, etc. |
| 76. | If you would like to hear a worded opinion, I would want AI to help and be a tool, like any other device, giving us insight that we wouldn't gather without. |
| 77. | AI provides a lot of possibilities in medical practice, from creating a diagnosis from scratch to analyzing certain medical scans/pictures (e.g., CT, histology, and so on). |
| 78. | Something that would be important to know is where the AI takes its information from, i.e., is it trustworthy or can it be manipulated (certain experiments from companies in social media show how some people can turn an AI into something that wasn't intended). |
| 79. | An important part of being able to use AI in certain evaluating processes is being able to critically weigh its final statements. |
| 80. | It always depends on how you use it. It could lead to ethical problems if you let the technology go its own way, but it doesn't really have to. I am not sure if it will change anything within ten years (given the "German speed"). |
| 81. | Great survey. It is definitely the topic that should be further integrated into medicine. In the end, the code runs more reliably than a human, especially with long working hours. |
| 82. | The amount of self-diagnosis from patients will rise, which brings another field of problems (e.g., wrong diagnosis and no trust in the physician/fear, etc. This aspect is not represented in the current questionnaire but is also important to think about. |
| 83. | Improve preparedness for AI; expertise is still irreplaceable. |
| 84. | In many departments, such as radiology, artificial intelligence is already used when creating images. Here, I find the application to be useful since such masses of data can no longer be processed by hand. |
| 85. | It would be very helpful to let AI do the documentation on its own. So, it is listening all the time and filtering out the important notes. |
| 86. | AI can take over tedious paperwork tasks, such as patient reports, etc. |
| 87. | I believe that legal and ethical problems already exist without the use of AI. |
| 88. | The role of AI in medicine wouldn't be a giving shitty diagnosis. The biggest benefit of AI is in administration work... |
| 89. | According to my personal experience in the first two years of university, I think that AI will be very important in helping the diagnosis of some rare diseases, interpreting some markers that can be confronted with billions of data, and finding particular diseases. |

| | |
|---|---|
| 90. | I like the question about what kind of subjects we should learn about AI in the curriculum. I strongly support only the two I ticked. I disagree with a subject on future perspectives of AI. It's just time-wasting to speculate on what we could do with it instead of trying to understand it more and actually create a new useful tool based on AI. |
| 91. | I personally would also be interested in the chances of using AI in the context of educational disparities. |
| 92. | Reliance on AI will produce incompetent doctors. |
| 93. | Our university should offer courses and training on AI for those who want to take them, and there should definitely be some subjects on AI. |
| 94. | The reason why I am so negative towards the implementation of AI in healthcare services is the fact that I see it as something that will replace human labor (as it will do with many other jobs). Therefore, I see it as a threat to my future career since it will probably be able to do everything I do (and maybe even more) but better than me. |
| 95. | We should use AI to manage the university. |
| 96. | AI will steal my job; I'm scared. |
| 97. | AI should be seen as a tool and not a substitute for physicians. In the end, it will be a human doctor to make a decision. You can never remove the human aspect of medicine. |
| 98. | Even though AI seems scary in some ways, we cannot withdraw developments that have already happened. Thus, we must know about its drawbacks as much as about its benefits. We need to learn to use it wisely and not exclusively let people deal with it who would use it for the wrong reasons. We must learn to integrate this new technology as well and as fast as possible in our daily lives. Otherwise, some other autocratic state will do, and we'll lose another battle besides the many we have already lost. I will be honest: I hate the idea of AI determining the decisions we make and the choices we take, but still, I think we can't bear not using it because emerging powers will do, and we must retain our competitiveness as well as our independence in reference to any sector of industry (medical treatment and science included). |
| 99. | In my opinion, AI will mostly affect the diagnosis process involving imaging techniques (CT, MRI, etc.), while surgery won't be affected as much. Patients will also use different AI tools to analyze their own symptoms. |
| 100. | I strongly support that AI should be a part of medical curriculum residency education programs... |
| 101. | I really do think this is a bad idea. |
| 102. | I'm currently in the midst of switching career paths from medicine to computer engineering, so my opinion might be slightly skewed. |
| 103. | I think these evaluations on how much or estimating effects are difficult to answer because if ethics and privacy are considered and accounted for, I think AI could make lots of improvements, but if ethics and privacy are just left by the wayside, I think it would have horrible effects. |
| 104. | I think that AI is going to really improve the healthcare physicians can provide to populations. Human error is still a very big problem in today's healthcare systems, and I think it is a moral obligation for physicians to use AI to give the best treatment and counseling to their patients. |
| 105. | I think, in the future, AI should definitely be used for diagnosis or surgical skills. However, I don't think it should ever be more valued than physicians or their opinions. |
| 106. | I think we will need to adapt AI to clinics. It's not something we should be afraid of. Contrarily, it will make doctors more comfortable by missing fewer diagnoses. |
| 107. | It's scary. The physician will do just the last check if the AI has done a good job. |
| 108. | The potential efficiency of AI cannot be analyzed through only its raw ability. It is influenced by many other factors, such as economic factors, political factors, and others. |
| 109. | I hope it doesn't take away jobs and people start solely depending on AI for diagnosis, for I believe medicine needs the human element. |
| 110. | The moment AI somehow gains self-consciousness, humanity is doomed. |
| 111. | AI makes life easier for doctors, from directing an approach to thinking around a diagnosis. |
| 112. | I believe AI can work as a helpful tool, especially for screening, but it shouldn't be used as gospel for diagnosis. |
| 113. | Let's talk more about using AI to solve economic and health records problems. |
| 114. | I think people need to be aware of the fallibility of AI and treat it as a consult rather than a definitive answer, and remember the final responsibility is with the doctor/themselves. |
| 115. | Let's not forget that AI doesn't have emotion; emotions and empathy are some of the characteristics of a good doctor. |
| 116. | Complete transition to exams done with computers in university under supervision. Sweden has transitioned into that, and other countries in Scandinavia. The pen-book process is an almost old method now for exams and tests. |
| 117. | (Practical) implementation of novel technologies is always difficult in (routine) healthcare processes. I wonder how easily AI will be available. |
| 118. | The use of AI in medicine disregards the humanistic aspect of medicine and may lead to less critical thinking, problem-solving, and team dynamics. I don't think it has a place in medicine except when considering patient safety and weighing the pros and cons in medical decision-making. |
| 119. | I would like to disclose a bias since I have personally worked at a company that develops large language models, and I currently work in a company that develops large language models for medical use. |
| 120. | We should not depend on AI to perform as we should as physicians, but it would be helpful for all of us. |
| 121. | Using artificial intelligence as a diagnostic tool, not as the actual diagnosis. |
| 122. | I think AI could have a place in medical study, but we can't forget how urgent medical students really need reasoning in studying various cases and materials related to medicine. |
| 123. | A physician's experience should always be valued, and the use of AI should be a complement to your logical reasoning. |
| 124. | I hadn't heard much about artificial intelligence, but answering this survey made me very curious to learn more about it. I believe that artificial intelligence, like any other technological device, has some positive aspects and, of course, some negative aspects. Thank you very much for the survey. |
| 125. | We need more education, events, etc., about AI. |
| 126. | The math is most important. Greatest fear: same treatment as with stats: executing code and algorithms that no one understands, even declaring knowledge as "unnecessary". Basic understanding is key, not heuristics. |
| 127. | Germany really refuses anything that is new. I think it is going to be the last country to start using AI in medicine. |
| 128. | 1. Protection of personal information is more important than training AI. 2. When we come to a point where AI is trained with AI-generated information, AI will be useless. |
| 129. | Preserve the health care worker's job. |
| 130. | I think it's not more important than focusing on improving knowledge and medical skills. |
| 131. | Despite believing that AI will bring great improvements to healthcare, I think one of the most important factors is the public's acceptance, which I believe will be low in the first years (for example, when people start to hear their diagnoses were made by a computer, etc.). |
| 132. | I think AI will be important to medicine. However, physicians need to be careful about how to use it and what decisions to make with the results of AI. What can be progress in one way can be a big problem in another. |

| | |
|---|---|
| 133. | AI is extremely helpful in gaining daily knowledge. |
| 134. | AI uses surgery diagnosis; everything should be included in the schedule of studies. |
| 135. | I believe in the fact that nothing is perfect in this world, so there might be some disadvantages, too. AI never replaces a physician... |
| 136. | The AI is still not totally qualified as I don't think it can properly address the psychology of a person. |
| 137. | I believe that greater incorporation of AI in medicine is necessary and inevitable. A balance must be struck with its use; it should be used as a tool to confirm diagnoses. If it is used as the first line of knowledge, a physician may feel less inclined to look outside of that recommendation. On the contrary, early use of AI to help guide a physician as to which medical disorders are most likely may help improve efficiency. |
| 138. | Most of my family looks for medicine on Google or AI-based websites, which leads to resistance and unproductive utilization of drugs. |
| 139. | I guess AI is good, but it becomes a problem when it takes on completely the clinician's work. |
| 140. | I wish to change all old practical tests and experiments in physiology and biochemistry with current advanced equipment... |
| 141. | As an MBBS, I would love to have classes on AI and its application on AI. It will give me an Edge against others and will also help me to treat my patients better, which will bring fulfillment and happiness to me. I strongly recommend adding courses on AI to the curriculum. |
| 142. | Even though I agree learning about artificial intelligence is important, where would you find the curriculum time to implement these teaching sessions when medical school is already saturated with learning activities? |
| 143. | We aren't ready for AI in medicine yet; we haven't, and we need more general overall improvement. I know AI is useful on its own, but the rate at which it's evolving is alarming. It might displace people from their jobs. |
| 144. | Artificial intelligence is very good if used positively. It shouldn't be abused or bastardized. |
| 145. | I think applying AI to medicine can be very positive and bring many improvements and develop both the medical world and society as a whole. However, we must first regulate, legislate, and educate in order to be aware of what exactly we are opening our doors to. |
| 146. | I feel like AI will make our work pointless. |
| 147. | I think a huge investment is made in AI. It will completely take the task of humans and increase unemployment, conflict, and poverty among nations. The jobless number will be increasing. The only IT computer field those who have knowledge about coding-decoding programming will only live in this world. This world will be cruel to the simple population. AI is a positive development, but we should use it in a better way. |
| 148. | While I don't think AI will improve the efficiency of healthcare processes in the next ten years, I think it will in the next 20 years. I think initially, it may cause more problems (in similar ways that other technologies have before) before it becomes more streamlined and efficient. |
| 149.<br>150. | Terrifying what AI will do to fulfillment, wealth equality, and the fabric of society.<br>The frequency with which I see my first-year peers turn to and rely on generative AI like ChatGPT to research and learn about new conditions and other course content deeply distresses me due to my concerns about both the AI's accuracy and my future colleagues' apparent lack of desire to ratify their findings with multiple (trusted) sources. I am deeply worried that our communities will come to be serviced by physicians who lack the curiosity, knowledge, and independence of thought required to meaningfully and reliably apply modern evidence-based medicine, and while this may prove to be an overreaction, I would prefer excessive caution over blindly worshipping AI. |
| 151. | In my opinion, it is important to discuss how AI can not only be incorporated into medicine but also regulated. There should be guidelines in what situations and to what extent AI should be used in medicine and scientific research. |
| 152. | The human doctor is not replaceable through AI. |
| 153. | Have a look at ETH's curriculum. Especially in the second and third years of our bachelor's, they have integrated many more technical aspects than any other University in Switzerland (in studying human medicine). |
| 154. | I believe that AI might have a much bigger impact on the efficiency of our healthcare system by providing tools that minimize automatable tasks, e.g., automatically generated physician's letters rather than by supporting the diagnostic process. |
| 155. | As a student in a 3rd world country (South Africa), I feel we will be behind the rest of the world when it comes to incorporating AI into our practice. We are still writing paper charts and have not even gone electric in that respect. But I wish to learn more about AI so that I, as a future physician, can remain competitive globally. |
| 156. | I would like to be educated on the potential AI tools that can be used to study in med school. |
| 157. | I think it will be very good for patient outcomes and cost but not good for physicians. |
| 158. | Teaching AI, as mentioned above, should become a central part of medical studies. |
| 159. | AI might also endanger the needs of physicians. |
| 160. | AI is dangerous if it's made to make a diagnosis. We, as future physicians, shall always be able to work without any AI tool. As long as the AI calculates some scores and helps with the unnecessary admin work, it's okay, but going any further than this is dangerous. And what happens if we get to the point where we, as physicians, aren't able to make a diagnosis ourselves if we are completely dependent on AI? Who is responsible for every misdiagnosis? The programmer of the AI or the physician who isn't able to diagnose himself, who has been taught to trust the AI and therefore has never learned how to make a diagnosis? |
| 161. | AI is on the rise, without a doubt. What worries me is that our empathy and focus on the patient is currently getting worse - we are understaffed and are so busy with other things that patient care is often secondary. I am worried that with AI doing a certain part of our job, such as triage, we will lose even more patient contact and that everything will be dehumanized - like a factory! Doctors won't speak with patients as often, etc. They will miss more subtexts than they do now, and we will rely too much on technology than on our training, and patient care will get worse. It's always a question of benefit, but sometimes, maybe focus more on what will be lost due to AI than gain. Though most people would disagree and say if patients get the right care, they won't care about interhuman conversations as well as empathy, etc. I think it's quite the contrary. |
| 162. | Medical schools need to teach their students how to use AI for coding when they conduct research. |
| 163. | I think it has a lot of use in imaging and investigations but not in history or exams. |
| 164. | We have to think about how patients can also use AI to diagnose themselves, which could create a conflict when physicians don't agree with the findings and examinations but the patient is convinced. |
| 165. | AI implementation in the current healthcare system will probably represent the limit to its widespread use. It seems that the Quebec government and healthcare system are very slow at adopting new technologies and making them universal use. I'm quite convinced that AI is going to be used in the private healthcare system much faster than in the public system. |
| 166. | AI should be a tool as much as a stethoscope is a tool in medicine. It should guide diagnosis and management without ever making the final call. A stethoscope never makes the diagnosis of a heart murmur; neither should AI. It is important that a human remains the final decision maker, as it is impossible to hold a machine accountable. |
| 167. | I think there is a fairly big dilemma around the confidentiality of information we share about patients. At this moment, if you share information about a patient on ChatGPT, he will keep it in memory, and in theory, someone could figure out a way to retrieve this information. I think it would be really important to create an AI based on ChatGPT for medical purposes to avoid confidentiality dilemmas. |
| 168. | Artificial intelligence does not and perhaps never will replace the clinical and human skills that are at the heart of the provision of good medicine. |
| 169. | It would be great to have an inter-university curriculum on AI so that medical students from all European countries have a common knowledge base (as opposed to everyone having their own curriculum) -> more resources budget leads to a better program rather than every uni having to tackle the subject on their own. |
| 170. | Instead of investing study time in AI, it would be better if someone solved the problem that every medicine has to do hours of paperwork every day - if we would spend less time on that, we could be more competitive! |
| 171. | As far as we need AI for the future, it should not replace physicians. AI and physicians should go hand in hand. |
| 172. | Collaboration between machines and humans is key in future healthcare. |
| 173. | I just think that regardless of AI systems being proficient, if we can't get doctors and healthcare professionals to change their ideas and values, it's never going to be seen in a positive light. There are also concerns about privacy and malfunctions, which I think would require rigorous research. |

| | |
|---|---|
| 174. | I'm afraid about the valorization of medical careers in case AI is implemented in an irresponsible way. |
| 175. | I would like to add that with enough research and discussion about this with current and future physicians, a reliable and accurate decision can be reached on whether AI will be suitable for use in the future. |
| 176. | AI in medicine is the future. |
| 177. | AI needs to assist physicians, not do the work for them. |
| 178. | It would be great as a tool available for physicians, but if not regulated, they can definitely substitute many doctors... |
| 179. | AI can be preferred but not over a physician's decision. |
| 180. | It is a potentially useful tool, but legal issues will bog it down immensely, similar to Tesla's driving mode, so I don't expect much in the next decade. Maybe in a longer period of time, we could expect more integration. |
| 181. | People's trust in AI for medical diagnosis should be improved because a great number of patients think that a physician who uses AI for diagnosis isn't capable enough of making it by itself. |
| 182. | AI can be a useful resource, but just that. |
| 183. | I guess my thoughts on AI are rather negative; I mean, it's a good start as a tool for learning but not for doing your job for you... Maybe as a second opinion for your diagnosis. |
| 184. | I think AI can be useful to physicians as a way to help come up with more diagnoses, but it shouldn't be used as a replacement or a crutch. |
| 185. | Given the importance of a good anamnesis for diagnosis, I think that artificial intelligence would be a "plus" that can help in certain cases, but it shouldn't be the main tool that the doctor has at his disposal and put when answering the item "I think working with AI as a physician is necessary to stay competitive". I think that a doctor could end up running the risk of "relaxing" and relying too much on the work of AI instead of his own knowledge. |
| 186. | In my opinion, AI will help with diagnosis and other practical things, but the profession will never be lost due to the need for a human dimension that is complementary to care. I don't think artificial intelligence can replace emotions and affection, which can be as curative as medicine. |
| 187. | AI has a good impact, making it easier to find information. |
| 188. | I think AI is too much of a powerful tool to neglect. |
| 189. | Generally, AI is not at the level of being sufficient to act alone in medicine. It can be used as a variable tool, but still, it is just a tool. |
| 190. | I want to learn AI prompting and usage in the medical line so that my efficiency gets boosted. However, I am not able to find a good platform to learn about AI applications in daily life. |
| 191. | In my point of view, I would rather prefer the old conservative way of diagnosis. But generally, we see that either physicians or the AI is held responsible for misdiagnosis when they do it alone. But if they combine both their knowledge, the chances of error might reduce. |
| 192. | AI should be used wisely. |
| 193. | Regarding the impact of AI on medicine in the medium and short term, what could we expect? |
| 194. | AI does not have intelligence on its own. It's really, really important for it to be a little over-sensitive so doctors can think about a diagnosis. If it were diagnosed, physicians would just believe it and leave many people without a diagnosis (due to overly trusting the AI). |
| 195. | AI is here to stay, and it will change the actual paradigm of our society. |
| 196. | I believe AI can really help with teaching in creative ways. |
| 197. | AI can be used as a great tool for second opinions, which may improve the diagnosis process. However, the only trustworthy source should be the physician's knowledge rather than a computer since it has made mistakes before. |
| 198. | I think we should also focus on what it means for us when diagnosing illnesses is less part of our job than appropriate interaction with our patients. |
| 199. | I personally would not like to be treated just by AI, without a person's at least "supervision". Do we, as future healthcare professionals, decrease our cognitive/manual competencies by passing our work to an electricity-driven, Wi-Fi-dependent AI? |
| 200. | For the use of AI in medicine, it's necessary to teach doctors about the limitations of AI. For example, the data AI is using is often biased. I see it as a new technology with potential and risks. But it's not going to go away, so we have to learn to use it accordingly. |
| 201. | I am a bit scared of AI. |
| 202. | Waiting for more in the future of artificial intelligence. |
| 203. | There are some elective courses at our university that talk about AI, but I'm not sure if it's included in the main curriculum. |
| 204. | I think it is very important to work to ensure that AI is no longer seen as a threat to physicians' employment but rather that it is recognized as a tool that will allow us to achieve excellent medical practice in a more efficient way. |
| 205. | Having knowledge of the use of artificial intelligence is necessary to update medical innovations. |
| 206. | I guess the use of AI is very dependent on the medical field (i.e., different possibilities in dermatology than in surgery). |
| 207. | I am concerned that AI would reduce the medical diagnostic and problem-solving abilities of medical professionals in the long run. As such, medical professionals should not be completely dependent on AI. |
| 208. | I strongly support that AI should be a part of the medical curriculum, not only in elective classes or seminars, but should be in the core structure. |
| 209. | I am currently working on research involving ML (LLMs) for emergency medicine uses. However, I don't know much about the coding and algorithm creation process (I am learning). We have an AI in medicine elective, which is optional currently, but no actual course lectures on AI. |
| 210. | I think there is a great benefit to using AI to improve inefficiencies in our current medical system (i.e., chart dictation, amalgamation of medical records, creating a holistic treatment plan with recommendations from new research), but these benefits need to be clearly monitored and regulated for bias and healthcare professionals (from all professions) should be involved in all levels development of this AI and not just rely on whatever big tech companies develop. |
| 211. | There is no need for teaching at this stage. |
| 212. | I think there are a lot of areas in which AI could be used to improve healthcare, but I feel like I have been told that it will take away job opportunities, and I am unsure of how true that is. |
| 213. | I can't say I feel prepared because I would probably be part of the generation that had no exposure to AI during my studies, but I would likely be part of the generation where it is tested on/piloted or introduced. It may assist in the healthcare burden we have, but with every radical change comes its implications/ host of problems. |
| 214. | AI, I believe, is a natural progression for us humans as it will help us greatly, but we will still need humans in order to manage AI maintenance and correct any problems. In other words, we will support the AI. |
| 215. | More lectures on artificial intelligence are needed. |
| 216. | Artificial intelligence will be the future of medicine. |
| 217. | I do believe that AI does have many aspects that are helpful in assisting professionals. We have seen this already with many business operations. However, I believe that, like what we see in business, AI is a tool to assist in operational aspects that lead to decision-making. But AI itself does not necessarily make the decision or diagnosis. Physicians make decisions with their expert clinical knowledge, but AI can help with the organization of charts, linking variables, and during patient interviews. |
| 218. | My thesis project was related to a chatbot used in a digital health application. |

| | |
|---|---|
| 219. | I think AI has a huge potential to be a transformative tool in medicine and medical practice; however, this depends entirely on the legal and social frameworks upon which it is implemented. At the moment, I do not have high hopes that the use of AI will be done in a way that is ethical, protective of human rights and freedoms, and promotes/advances human health/wellbeing. |
| 220. | AI is an unknown abyss for now. |
| 221. | The introduction of AI into the medical curriculum should begin in the pre-clinical or pre-medical years. Also, seasoned medical practitioners should be encouraged to take courses on AI as it relates to medicine. |
| 222. | My problem with quality healthcare in Africa, particularly in Sub-Saharan Africa, is that it's too expensive, and it's only affordable by the elites, leaving the poor masses almost completely out. Won't AI be for just the rich? |
| 223. | I think AI could be an excellent asset in healthcare if implemented properly. |
| 224. | I believe it's being implemented way too quickly. It must be further developed and understood by its developers. There must be proper education and training around how AI works and how to use it effectively. There must be adequate and thorough legal and ethical measures in place around using AI in medicine. None of these have been accomplished, and I feel the rate at which it's being integrated is completely negligent and irresponsible. |
| 225. | I think that, in general, AI technology is a great innovation with many advantages in medical practice. However, it may have some serious negative consequences on society in terms of over-reliance and missed diagnosis if not developed adequately to solve those potential problems. That will be especially dangerous to the future of humanity. |
| 226. | AI is the future, and since we will be the future doctors, we should have some workshops to teach us how to work with them so we can be better. |
| 227. | Academics should take a more serious approach towards AI. |
| 228. | I wish I had more time for AI research. |
| 229. | I feel students should be encouraged to integrate AI into completing tasks in the medical setting, along with peer-reviewed resources we are using. |
| 230. | Job security is a massive fear with regard to the implantation and introduction of AI in many workspaces, and this is no different way in medicine. |
| 231. | I believe that AI can be just as revolutionary in medicine as it is in other fields. However, more emphasis should be put on screening for, detecting, and addressing the legal and ethical issues surrounding its application in medicine. It's all about striking a balance between safety and the efficiency we so desperately want. |
| 232. | I want there to be more cross-discipline collaboration between the medical and technical faculties. We need to both understand our languages and thus make useful contributions and implementations to practical settings. |
| 233. | I worry about the data in AI not being representable and unbiased to be able to hold the role of a second opinion. |
| 234. | I am afraid AI will take over radiology and pathology jobs. |
| 235. | AI is very important for the development of medicine. I think it is important to know a little more about it. |
| 236. | More AI education is needed. |
| 237. | AI is the future, so more education. |
| 238. | More events related to AI are needed. |
| 239. | More AI lectures are needed. |
| 240. | I agree that the integration of AI into medical education and clinical practice has significantly boosted the productivity of medical students and physicians. |
| 241. | AI is a tool, and it will depend on the criteria and professional quality of those who use it to achieve the expected results. |
| 242. | AI, like ChatGPT, has helped me to study because, based on a textbook, it can apply an exam or explain things in a more simplified way. |
| 243. | AI should always function as a second-choice tool, never fully replacing a physician and his or her opinion. It gives a lot to think about how the use of AI in health care training is controlled, talking about academic dishonesty. |
| 244. | I would like to warn you about AI. Although it is an information-rich program, I think it is very important and valuable to emphasize that the physician is not a walking repertory of information. I believe that AI can provide a certain amount of support for a doubtful or difficult diagnosis. However, I believe that medical decisions cannot be based on AI responses. Especially considering epidemiological contexts. Even more so considering areas such as Tropical Medicine since (for example) it will never be possible to compare a Latin American patient contracting neurocysticercosis to a European patient. In summary, I believe that the issue of AI should be taken with caution and, as mentioned in the information section of the study, a way should be sought for AI to be an instrument that supports the development of medicine, medical education, and the good of humanity, rather than a universal "solvent" in medicine. |
| 245. | AI is a very interesting tool and will soon be indispensable for medical practice. |
| 246. | Definitely, in the future, a doctor who does not know how to use AI as a daily work tool will be left far behind at a competitive level with his peers. |
| 247. | I think it is important to move forward with technology because it makes us stronger, but I don't think we should get to a point where physicians rely solely on AIs to work on medical records. The facilities of AIs are creating generations that are increasingly incapable and over-dependent on AIs. Physicians cannot be dependent on something that can fail at any time (power outage, putting us in a drastic situation, or even human failure in creating these technologies) because it will be in those moments of greatest difficulty that the entire population will put their trust in us, and we cannot afford to have fewer competencies or knowledge because we have put our blindest trust in AIs. I like the idea of learning to integrate them into the profession and seeing them as a support, but making it clear that they are subject to error or unavailability at any time. Finally, I would like to remind future generations of physicians: let our brains be the most important thing in learning medicine. |
| 248. | I know it is a very useful tool, but I don't know how good it will be in decision-making. I prefer to make them personally and at my own discretion, but as a source of knowledge, it is very good. |
| 249. | AI, properly used, can be a great tool, not that it replaces the physician, but rather a support to improve the efficiency of the professional. |
| 250. | I believe that AI can help train physicians more interactively. |
| 251. | AI must be adapted as another digital tool. |
| 252. | At the end of the day, medicine is a humanistic career. We need humans to carry out this profession. However, AI could be of great help to create alternatives in the diagnosis or treatment of a patient, but not to deal with them. |
| 253. | I do not really know how AI can help in medicine to this day. |
| 254. | I believe that it would also be of great importance to establish limits in the use of AI because although it would facilitate many activities, it could also generate problems in learning. |
| 255. | I would like to learn more about AI. |
| 256. | AI is the future. |
| 257. | I would like to know more about the development of AI in the medical field with the new technology now. |
| 258. | I believe that AI is a necessary but very dangerous tool and must be properly controlled. |
| 259. | It would be good to integrate it into both university and hospital teaching. |

Grammar and spelling have been adjusted where appropriate. Translation into American English, if applicable, was carried out with DeepL.

### S3 Table. Subgroup analysis by gender.

| Item/Group | Median (IQR) | P-value | r |
|---|---|---|---|
| **Attitude towards medical studies** | - | - | - |
| What is your current general attitude toward your medical studies? (N=4,529) | | | |
| Female (N=2,587) | 4 (3-4) | <.001 | .035 |
| Male (N=1,942) | 4 (4-4) | | |
| **Perspectives towards AI in the medical profession** | - | - | - |
| What is your general attitude toward the application of artificial intelligence (AI) in medicine? (N=4,526) | | | |
| Female (N=2,587) | 4 (3-4) | .017 | .133 |
| Male (N=1,939) | 4 (3-4) | | |
| How do you estimate the effect of artificial intelligence (AI) on the efficiency of healthcare processes in the next 10 years? (N=4,520) | | | |
| Female (N=2,583) | 4 (4-5) | <.001 | .082 |
| Male (N=1,937) | 4 (4-5) | | |
| The use of artificial intelligence (AI) in medicine will increasingly lead to legal and ethical conflicts. (N=4,525) | | | |
| Female (N=2,585) | 4 (3-5) | .492 | .010 |
| Male (N=1,940) | 4 (3-5) | | |
| What is your view on the influence of artificial intelligence (AI) on the profession of physicians? AI will affect the everyday life of physicians in a way that is… (N=4,522) | | | |
| Female (N=2,584) | 4 (3-4) | <.001 | .101 |
| Male (N=1,938) | 4 (3-4) | | |
| How would you rate artificial intelligence (AI) software being available to physicians as a second opinion on medical issues? (N=4,516) | | | |
| Female (N=2,583) | 4 (3-4) | <.001 | .080 |
| Male (N=1,933) | 4 (3-4) | | |
| I think working with artificial intelligence (AI) as a physician is necessary to stay competitive. (N=4,526) | | | |
| Female (N=2,586) | 4 (3-4) | <.001 | .154 |
| Male (N=1,940) | 4 (3-4) | | |
| With my current knowledge, I feel sufficiently prepared to work with artificial intelligence (AI) in my future profession as a physician. (N=4,527) | | | |
| Female (N=2,587) | 2 (1-3) | <.001 | .146 |
| Male (N=1,940) | 2 (2-4) | | |
| **AI education and knowledge level** | - | - | - |
| I would like to have more teaching on artificial intelligence (AI) in medicine as part of my studies. (N=4,515) | | | |
| Female (N=2,578) | 4 (3-5) | <.001 | .087 |
| Male (N=1,937) | 4 (4-5) | | |
| As part of my studies, there are curricular events on artificial intelligence (AI) in medicine. (N=4,531) | | | |
| Female (N=2,588) | 1 (1-1) | .335 | .014 |
| Male (N=1,943) | 1 (1-1) | | |
| How would you rate your general knowledge of artificial intelligence (AI)? (N=4,535) | | | |
| Female (N=2,593) | 2 (2-2) | <.001 | .190 |
| Male (N=1,942) | 2 (2-3) | | |

The Mann-Whitney U-test was used to compare the subgroups. Significant p-values display higher Likert scale ranks by male respondents.

IQR, interquartile range.

### S4 Table. Subgroup analysis by age.

| Item/Group | Median (IQR) | P-value | r |
|---|---|---|---|
| **Attitude towards medical studies** | - | - | - |
| What is your current general attitude toward your medical studies? (N=4,554)<br>≤ 22 years of age (N=2,591)<br>> 22 years of age (N=1,963) | 4 (4-4)<br>4 (3-4) | .043* | .030 |
| **Perspectives towards AI in the medical profession** | - | - | - |
| What is your general attitude toward the application of artificial intelligence (AI) in medicine? (N=4,551)<br>≤ 22 years of age (N=2,588)<br>> 22 years of age (N=1,963) | 4 (3-4)<br>4 (3-4) | .890 | .002 |
| How do you estimate the effect of artificial intelligence (AI) on the efficiency of healthcare processes in the next 10 years? (N=4,545)<br>≤ 22 years of age (N=2,586)<br>> 22 years of age (N=1,959) | 4 (4-5)<br>4 (4-5) | .019† | .035 |
| The use of artificial intelligence (AI) in medicine will increasingly lead to legal and ethical conflicts. (N=4,550)<br>≤ 22 years of age (N=2,587)<br>> 22 years of age (N=1,963) | 4 (3-5)<br>4 (3-5) | <.001† | .081 |
| What is your view on the influence of artificial intelligence (AI) on the profession of physicians? AI will affect the everyday life of physicians in a way that is… (N=4,547)<br>≤ 22 years of age (N=2,584)<br>> 22 years of age (N=1,963) | 4 (3-4)<br>4 (3-4) | .022† | .034 |
| How would you rate artificial intelligence (AI) software being available to physicians as a second opinion on medical issues? (N=4,541)<br>≤ 22 years of age (N=2,581)<br>> 22 years of age (N=1,960) | 4 (3-4)<br>4 (3-4) | .699 | .006 |
| I think working with artificial intelligence (AI) as a physician is necessary to stay competitive. (N=4,552)<br>≤ 22 years of age (N=2,587)<br>> 22 years of age (N=1,965) | 4 (3-4)<br>4 (3-4) | <.001† | .052 |
| With my current knowledge, I feel sufficiently prepared to work with artificial intelligence (AI) in my future profession as a physician. (N=4,552)<br>≤ 22 years of age (N=2,587)<br>> 22 years of age (N=1,965) | 2 (2-3)<br>2 (2-3) | .390 | .013 |
| **AI education and knowledge level** | - | - | - |
| I would like to have more teaching on artificial intelligence (AI) in medicine as part of my studies. (N=4,541)<br>≤ 22 years of age (N=2,582)<br>> 22 years of age (N=1,959) | 4 (4-5)<br>4 (4-5) | .465 | .011 |
| As part of my studies, there are curricular events on artificial intelligence (AI) in medicine. (N=4,556)<br>≤ 22 years of age (N=2,590)<br>> 22 years of age (N=1,966) | 1 (1-1)<br>1 (1-1) | .087 | .025 |
| How would you rate your general knowledge of artificial intelligence (AI)? (N=4,560)<br>≤ 22 years of age (N=2,592)<br>> 22 years of age (N=1,968) | 2 (2-2)<br>2 (2-3) | <.001† | .071 |

The Mann-Whitney U-test was used to compare the subgroups.

* Higher Likert scale ranks by respondents below or equal to the median age of 22 years.

† Higher Likert scale ranks by respondents exceeding the median age of 22 years.

IQR, interquartile range.

### S5 Table. Subgroup analysis by study year.

| Item/Group | Median (IQR) | P-value | r |
|---|---|---|---|
| **Attitude towards medical studies** | - | - | - |
| What is your current general attitude toward your medical studies? (N=4,456) | | | |
| ≤ 3 academic years (N=2,272) | 4 (4-4) | <.001* | .067 |
| > 3 academic years (N=2,184) | 4 (3-4) | | |
| **Perspectives towards AI in the medical profession** | - | - | - |
| What is your general attitude toward the application of artificial intelligence (AI) in medicine? (N=4,453) | | | |
| ≤ 3 academic years (N=2,269) | 4 (3-4) | 033* | .032 |
| > 3 academic years (N=2,184) | 4 (3-4) | | |
| How do you estimate the effect of artificial intelligence (AI) on the efficiency of healthcare processes in the next 10 years? (N=4,450) | | | |
| ≤ 3 academic years (N=2,269) | 4 (4-5) | .533 | .009 |
| > 3 academic years (N=2,181) | 4 (4-5) | | |
| The use of artificial intelligence (AI) in medicine will increasingly lead to legal and ethical conflicts. (N=4,452) | | | |
| ≤ 3 academic years (N=2,268) | 4 (3-5) | .049† | .029 |
| > 3 academic years (N=2,184) | 4 (3-5) | | |
| What is your view on the influence of artificial intelligence (AI) on the profession of physicians? AI will affect the everyday life of physicians in a way that is… (N=4,451) | | | |
| ≤ 3 academic years (N=2,267) | 4 (3-4) | .082 | .026 |
| > 3 academic years (N=2,184) | 4 (3-4) | | |
| How would you rate artificial intelligence (AI) software being available to physicians as a second opinion on medical issues? (N=4,448) | | | |
| ≤ 3 academic years (N=2,267) | 4 (3-4) | .190 | .020 |
| > 3 academic years (N=2,181) | 4 (4-4) | | |
| I think working with artificial intelligence (AI) as a physician is necessary to stay competitive. (N=4,454) | | | |
| ≤ 3 academic years (N=2,269) | 4 (3-4) | .630 | .007 |
| > 3 academic years (N=2,185) | 4 (3-4) | | |
| With my current knowledge, I feel sufficiently prepared to work with artificial intelligence (AI) in my future profession as a physician. (N=4,454) | | | |
| ≤ 3 academic years (N=2,270) | 2 (2-3) | <.001* | .101 |
| > 3 academic years (N=2,184) | 2 (2-4) | | |
| **AI education and knowledge level** | - | - | - |
| I would like to have more teaching on artificial intelligence (AI) in medicine as part of my studies. (N=4,443) | | | |
| ≤ 3 academic years (N=2,266) | 4 (4-5) | <.001* | .050 |
| > 3 academic years (N=2,177) | 4 (3-5) | | |
| As part of my studies, there are curricular events on artificial intelligence (AI) in medicine. (N=4,460) | | | |
| ≤ 3 academic years (N=2,273) | 1 (1-1) | .253 | .017 |
| > 3 academic years (N=2,187) | 1 (1-1) | | |
| How would you rate your general knowledge of artificial intelligence (AI)? (N=4,464) | | | |
| ≤ 3 academic years (N=2,275) | 2 (2-2) | <.001† | .053 |
| > 3 academic years (N=2,189) | 2 (2-3) | | |

The Mann-Whitney U-test was used to compare the subgroups.

* Higher Likert scale ranks by respondents below or equal to the median of three academic years.

† Higher Likert scale ranks by respondents exceeding the median of three academic years.

IQR, interquartile range.

### S6 Table. Subgroup analysis by weekly use of technical devices.

| Item/Group | Median (IQR) | P-value | r |
|---|---|---|---|
| **Attitude towards medical studies** | - | - | - |
| What is your current general attitude toward your medical studies? (N=4,579) | | | |
| Technical devices used ≤ 3 (N=2.014) | 4 (3-4) | .036* | .031 |
| Technical devices used > 3 (N=2,565) | 4 (4-4) | | |
| **Perspectives towards AI in the medical profession** | - | - | - |
| What is your general attitude toward the application of artificial intelligence (AI) in medicine? (N=4,576) | | | |
| Technical devices used ≤ 3 (N=2,013) | 4 (3-4) | <.001† | .050 |
| Technical devices used > 3 (N=2,563) | 4 (3-4) | | |
| How do you estimate the effect of artificial intelligence (AI) on the efficiency of healthcare processes in the next 10 years? (N=4,570) | | | |
| Technical devices used ≤ 3 (N=2,009) | 4 (4-5) | <.001† | .076 |
| Technical devices used > 3 (N=2,561) | 4 (4-5) | | |
| The use of artificial intelligence (AI) in medicine will increasingly lead to legal and ethical conflicts. (N=4,575) | | | |
| Technical devices used ≤ 3 (N=2,014) | 4 (3-5) | <.001† | .084 |
| Technical devices used > 3 (N=2,561) | 4 (3-5) | | |
| What is your view on the influence of artificial intelligence (AI) on the profession of physicians? AI will affect the everyday life of physicians in a way that is… (N=4,571) | | | |
| Technical devices used ≤ 3 (N=2,009) | 4 (3-4) | <.001† | .079 |
| Technical devices used > 3 (N=2,562) | 4 (3-4) | | |
| How would you rate artificial intelligence (AI) software being available to physicians as a second opinion on medical issues? (N=4,565) | | | |
| Technical devices used ≤ 3 (N=2,007) | 4 (3-4) | <.001† | .073 |
| Technical devices used > 3 (N=2,558) | 4 (4-4) | | |
| I think working with artificial intelligence (AI) as a physician is necessary to stay competitive. (N=4,576) | | | |
| Technical devices used ≤ 3 (N=2,013) | 4 (3-4) | <.001† | .058 |
| Technical devices used > 3 (N=2,563) | 4 (3-4) | | |
| With my current knowledge, I feel sufficiently prepared to work with artificial intelligence (AI) in my future profession as a physician. (N=4,577) | | | |
| Technical devices used ≤ 3 (N=2,015) | 2 (2-3) | <.001† | .064 |
| Technical devices used > 3 (N=2,562) | 2 (2-3) | | |
| **AI education and knowledge level** | - | - | - |
| I would like to have more teaching on artificial intelligence (AI) in medicine as part of my studies. (N=4,565) | | | |
| Technical devices used ≤ 3 (N=2,007) | 4 (3-5) | <.001* | .080 |
| Technical devices used > 3 (N=2,558) | 4 (4-5) | | |
| As part of my studies, there are curricular events on artificial intelligence (AI) in medicine. (N=4,581) | | | |
| Technical devices used ≤ 3 (N=2,019) | 1 (1-1) | .786 | .004 |
| Technical devices used > 3 (N=2,562) | 1 (1-1) | | |
| How would you rate your general knowledge of artificial intelligence (AI)? (N=4,585) | | | |
| Technical devices used ≤ 3 (N=2,020) | 2 (2-2) | <.001† | .094 |
| Technical devices used > 3 (N=2,565) | 2 (2-3) | | |

The Mann-Whitney U-test was used to compare the subgroups.

* Higher Likert scale ranks by respondents below or equal to the median number of devices used per week.

† Higher Likert scale ranks by respondents exceeding the median number of devices used per week.

IQR, interquartile range.

### S7 Table. Subgroup analysis by previous coding experience.

| Item/Group | Median (IQR) | P-value | r |
|---|---|---|---|
| **Attitude towards medical studies** | - | - | - |
| What is your current general attitude toward your medical studies? (N=4,576) | | | |
| Have coded (N=908) | 4 (3-4) | .205 | .019 |
| Have not coded (N=3,668) | 4 (3-4) | | |
| **Perspectives towards AI in the medical profession** | - | - | - |
| What is your general attitude toward the application of artificial intelligence (AI) in medicine? (N=4,573) | | | |
| Have coded (N=908) | 4 (3-5) | <.001 | .102 |
| Have not coded (N=3,665) | 4 (3-4) | | |
| How do you estimate the effect of artificial intelligence (AI) on the efficiency of healthcare processes in the next 10 years? (N=4,567) | | | |
| Have coded (N=909) | 4 (4-5) | <.001 | .059 |
| Have not coded (N=3658) | 4 (4-5) | | |
| The use of artificial intelligence (AI) in medicine will increasingly lead to legal and ethical conflicts. (N=4572) | | | |
| Have coded (N=910) | 4 (4-5) | <.001 | .102 |
| Have not coded (N=3,662) | 4 (3-5) | | |
| What is your view on the influence of artificial intelligence (AI) on the profession of physicians? AI will affect the everyday life of physicians in a way that is… (N=4,568) | | | |
| Have coded (N=910) | 4 (4-4) | <.001 | .080 |
| Have not coded (N=3,658) | 4 (3-4) | | |
| How would you rate artificial intelligence (AI) software being available to physicians as a second opinion on medical issues? (N=4,562) | | | |
| Have coded (N=909) | 4 (3-4) | .035 | .031 |
| Have not coded (N=3,653) | 4 (3-4) | | |
| I think working with artificial intelligence (AI) as a physician is necessary to stay competitive. (N=4,573) | | | |
| Have coded (N=909) | 4 (3-5) | <.001 | .079 |
| Have not coded (N=3,664) | 4 (3-4) | | |
| With my current knowledge, I feel sufficiently prepared to work with artificial intelligence (AI) in my future profession as a physician. (N=4,574) | | | |
| Have coded (N=910) | 2 (2-4) | <.001 | .087 |
| Have not coded (N=3,664) | 2 (2-3) | | |
| **AI education and knowledge level** | - | - | - |
| I would like to have more teaching on artificial intelligence (AI) in medicine as part of my studies. (N=4,562) | | | |
| Have coded (N=909) | 4 (4-5) | <.001 | .082 |
| Have not coded (N=3,653) | 4 (4-5) | | |
| As part of my studies, there are curricular events on artificial intelligence (AI) in medicine. (N=4,578) | | | |
| Have coded (N=910) | 1 (1-2) | <.001 | .072 |
| Have not coded (N=3,668) | 1 (1-1) | | |
| How would you rate your general knowledge of artificial intelligence (AI)? (N=4,582) | | | |
| Have coded (N=910) | 2 (2-3) | <.001 | .214 |
| Have not coded (N=3,672) | 2 (2-2) | | |

The Mann-Whitney U-test was used to compare the subgroups. Significant p-values display higher Likert scale ranks by respondents who have already programmed code.

IQR, interquartile range.

### S8 Table. Subgroup analysis by AI knowledge.

| Item/Group | Median (IQR) | P-value | r |
|---|---|---|---|
| **Attitude towards medical studies** | - | - | - |
| What is your current general attitude toward your medical studies? (N=4,577) | | | |
| No or little AI knowledge (N=3,444) | 4 (3-4) | <.001 | .078 |
| Good or expert AI knowledge (N=1,133) | 4 (4-4) | | |
| **Perspectives towards AI in the medical profession** | - | - | - |
| What is your general attitude toward the application of artificial intelligence (AI) in medicine? (N=4,574) | | | |
| No or little AI knowledge (N=3,443) | 4 (3-4) | <.001 | .170 |
| Good or expert AI knowledge (N=1,131) | 4 (4-5) | | |
| How do you estimate the effect of artificial intelligence (AI) on the efficiency of healthcare processes in the next 10 years? (N=4,568) | | | |
| No or little AI knowledge (N=3,440) | 4 (4-5) | <.001 | .125 |
| Good or expert AI knowledge (N=1,128) | 4 (4-5) | | |
| The use of artificial intelligence (AI) in medicine will increasingly lead to legal and ethical conflicts. (N=4,573) | | | |
| No or little AI knowledge (N=3,443) | 4 (3-5) | .804 | .004 |
| Good or expert AI knowledge (N=1,130) | 4 (3-5) | | |
| What is your view on the influence of artificial intelligence (AI) on the profession of physicians? AI will affect the everyday life of physicians in a way that is… (N=4,569) | | | |
| No or little AI knowledge (N=3,441) | 4 (3-4) | <.001 | .124 |
| Good or expert AI knowledge (N=1,128) | 4 (4-4) | | |
| How would you rate artificial intelligence (AI) software being available to physicians as a second opinion on medical issues? (N=4,563) | | | |
| No or little AI knowledge (N=3,437) | 4 (3-4) | <.001 | .077 |
| Good or expert AI knowledge (N=1,126) | 4 (3-4) | | |
| I think working with artificial intelligence (AI) as a physician is necessary to stay competitive. (N=4,574) | | | |
| No or little AI knowledge (N=3,443) | 4 (3-4) | <.001 | .162 |
| Good or expert AI knowledge (N=1,131) | 4 (3-5) | | |
| With my current knowledge, I feel sufficiently prepared to work with artificial intelligence (AI) in my future profession as a physician. (N=4,575) | | | |
| No or little AI knowledge (N=3,444) | 2 (1-3) | <.001 | .269 |
| Good or expert AI knowledge (N=1,131) | 3 (2-4) | | |
| **AI education and knowledge level** | - | - | - |
| I would like to have more teaching on artificial intelligence (AI) in medicine as part of my studies. (N=4,563) | | | |
| No or little AI knowledge (N=3,435) | 4 (3-5) | <.001 | .146 |
| Good or expert AI knowledge (N=1,128) | 4 (4-5) | | |
| As part of my studies, there are curricular events on artificial intelligence (AI) in medicine. (N=4,580) | | | |
| No or little AI knowledge (N=3,448) | 1 (1-1) | <.001 | .148 |
| Good or expert AI knowledge (N=1,132) | 1 (1-2) | | |
| How would you rate your general knowledge of artificial intelligence (AI)? (N=4,585) | | | |
| No or little AI knowledge (N=3,451) | 2 (2-2) | - | - |
| Good or expert AI knowledge (N=1,134) | 3 (3-3) | | |

The Mann-Whitney U-test was used to compare the subgroups. Significant p-values display higher Likert scale ranks by respondents who rated their AI knowledge as good or expert. IQR, interquartile range.

**S9 Table. Subgroup analysis by students who reported curricular events on AI in medicine of any duration versus those who indicated no curricular events.**

| Item/Group | Median (IQR) | P-value | r |
|---|---|---|---|
| **Attitude towards medical studies** | - | - | - |
| What is your current general attitude toward your medical studies? (N=4,574) | | | |
| No AI in the medical curriculum (N=3,493) | 4 (4-4) | .693 | .006 |
| AI in the medical curriculum (N=1,081) | 4 (3-4) | | |
| **Perspectives towards AI in the medical profession** | - | - | - |
| What is your general attitude toward the application of artificial intelligence (AI) in medicine? (N=4,572) | | | |
| No AI in the medical curriculum (N=3,493) | 4 (3-4) | .418 | .012 |
| AI in the medical curriculum (N=1,079) | 4 (3-4) | | |
| How do you estimate the effect of artificial intelligence (AI) on the efficiency of healthcare processes in the next 10 years? (N=4,565) | | | |
| No AI in the medical curriculum (N=3,489) | 4 (4-5) | .837 | .003 |
| AI in the medical curriculum (N=1,076) | 4 (4-5) | | |
| The use of artificial intelligence (AI) in medicine will increasingly lead to legal and ethical conflicts. (N=4,570) | | | |
| No AI in the medical curriculum (N=3,492) | 4 (3-5) | .983 | .000 |
| AI in the medical curriculum (N=1,078) | 4 (3-5) | | |
| What is your view on the influence of artificial intelligence (AI) on the profession of physicians? AI will affect the everyday life of physicians in a way that is… (N=4,566) | | | |
| No AI in the medical curriculum (N=3,489) | 4 (3-4) | .725 | .005 |
| AI in the medical curriculum (N=1,077) | 4 (3-4) | | |
| How would you rate artificial intelligence (AI) software being available to physicians as a second opinion on medical issues? (N=4,561) | | | |
| No AI in the medical curriculum (N=3,487) | 4 (3-4) | .075 | .026 |
| AI in the medical curriculum (N=1,074) | 4 (4-4) | | |
| I think working with artificial intelligence (AI) as a physician is necessary to stay competitive. (N=4,571) | | | |
| No AI in the medical curriculum (N=3,493) | 4 (3-4) | .118 | .023 |
| AI in the medical curriculum (N=1,078) | 4 (3-4) | | |
| With my current knowledge, I feel sufficiently prepared to work with artificial intelligence (AI) in my future profession as a physician. (N=4,572) | | | |
| No AI in the medical curriculum (N=3,493) | 2 (1-3) | <.001† | .201 |
| AI in the medical curriculum (N=1,079) | 3 (2-4) | | |
| **AI education and knowledge level** | - | - | - |
| I would like to have more teaching on artificial intelligence (AI) in medicine as part of my studies. (N=4,560) | | | |
| No AI in the medical curriculum (N=3,487) | 4 (4-5) | <.001* | .054 |
| AI in the medical curriculum (N=1,073) | 4 (3-5) | | |
| As part of my studies, there are curricular events on artificial intelligence (AI) in medicine. (N=4,581) | | | |
| No AI in the medical curriculum (N=3,497) | 1 (1-1) | - | - |
| AI in the medical curriculum (N=1,084) | 2 (2-2) | | |
| How would you rate your general knowledge of artificial intelligence (AI)? (N=4,580) | | | |
| No AI in the medical curriculum (N=3,496) | 2 (2-2) | <.001† | .137 |
| AI in the medical curriculum (N=1,084) | 2 (2-3) | | |

The Mann-Whitney U-test was used to compare the subgroups.

* Higher Likert scale ranks by respondents reporting no curricular events on medicine.

† Higher Likert scale ranks by respondents reporting curricular events on AI in medicine of any duration.

IQR, interquartile range.

### S10 Table. STROBE Statement—Checklist of items that should be included in reports of cross-sectional studies

| | Item No | Recommendation | In-text reference |
|---|---|---|---|
| **Title and abstract** | 1 | (*a*) Indicate the study's design with a commonly used term in the title or the abstract | Page 1, lines 2-3 |
| | | (*b*) Provide in the abstract an informative and balanced summary of what was done and what was found | Page 2, lines 35-49 |
| **Introduction** | | | |
| Background/rationale | 2 | Explain the scientific background and rationale for the investigation being reported | Pages 4-5, lines 74-91 |
| Objectives | 3 | State specific objectives, including any prespecified hypotheses | Page 5, lines 91-97 |
| **Methods** | | | |
| Study design | 4 | Present key elements of study design early in the paper | Page 6, lines 107-111 |
| Setting | 5 | Describe the setting, locations, and relevant dates, including periods of recruitment, exposure, follow-up, and data collection | Page 7, lines 131-147/S9 Table |
| Participants | 6 | (*a*) Give the eligibility criteria, and the sources and methods of selection of participants | Page 7, lines 150-156/S9 Table |
| Variables | 7 | Clearly define all outcomes, exposures, predictors, potential confounders, and effect modifiers. Give diagnostic criteria, if applicable | Page 6, lines 107-130/ pages 8-9, lines 159-184 |
| Data sources/ measurement | 8* | For each variable of interest, give sources of data and details of methods of assessment (measurement). Describe comparability of assessment methods if there is more than one group | Page 6, lines 111-130 |
| Bias | 9 | Describe any efforts to address potential sources of bias | Pages 6-7, lines 124-141 |
| Study size | 10 | Explain how the study size was arrived at | Page 7, lines 144-147 |
| Quantitative variables | 11 | Explain how quantitative variables were handled in the analyses. If applicable, describe which groupings were chosen and why | Page 8, lines 161-164/172-182 |
| Statistical methods | 12 | (*a*) Describe all statistical methods, including those used to control for confounding | Pages 8-9, lines 159-184 |
| | | (*b*) Describe any methods used to examine subgroups and interactions | Pages 8-9, lines 172-184 |
| | | (*c*) Explain how missing data were addressed | Pages 7, lines 155-156 |
| | | (*d*) If applicable, describe analytical methods taking account of sampling strategy | N/A |
| | | (*e*) Describe any sensitivity analyses | N/A |
| **Results** | | | |
| Participants | 13* | (a) Report numbers of individuals at each stage of study—eg numbers potentially eligible, examined for eligibility, confirmed eligible, included in the study, completing follow-up, and analysed | Pages 9-10, lines 206-213 |
| | | (b) Give reasons for non-participation at each stage | Page 10, lines 208-213 |
| | | (c) Consider use of a flow diagram | N/A |
| Descriptive data | 14* | (a) Give characteristics of study participants (eg demographic, clinical, social) and information on exposures and potential confounders | Page 10, lines 211-227/Table 1 |
| | | (b) Indicate number of participants with missing data for each variable of interest | Table 1/Table 2/S3-S9 Tables |
| Outcome data | 15* | Report numbers of outcome events or summary measures | Table 1/Table 2/S3-S9 Tables |
| Main results | 16 | (*a*) Give unadjusted estimates and, if applicable, confounder-adjusted estimates and their precision (eg, 95% confidence interval). Make clear which confounders were adjusted for and why they were included | Table 1/Table 2/S3-S9 Tables |
| | | (*b*) Report category boundaries when continuous variables were categorized | S4-S6 Tables |
| | | (*c*) If relevant, consider translating estimates of relative risk into absolute risk for a meaningful time period | N/A |
| Other analyses | 17 | Report other analyses done—eg analyses of subgroups and interactions, and sensitivity analyses | Page 20, lines 294-319/Table 2 |
| **Discussion** | | | |
| Key results | 18 | Summarise key results with reference to study objectives | Page 21, lines 322-331 |
| Limitations | 19 | Discuss limitations of the study, taking into account sources of potential bias or imprecision. Discuss both direction and magnitude of any potential bias | Page 23, lines 395-396/page 24, lines 406-418 |
| Interpretation | 20 | Give a cautious overall interpretation of results considering objectives, limitations, multiplicity of analyses, results from similar studies, and other relevant evidence | Pages 21-24, lines 332-418 |
| Generalisability | 21 | Discuss the generalisability (external validity) of the study results | Page 23, lines 395-396/page 24, lines 406-418 |
| **Other information** | | | |
| Funding | 22 | Give the source of funding and the role of the funders for the present study and, if applicable, for the original study on which the present article is based | Financial disclosure statement |

*Give information separately for exposed and unexposed groups.
